# The effects of COVID-19 on cognitive performance in a community-based cohort: A COVID Symptom Study Biobank observational study

**DOI:** 10.1101/2023.03.14.23287211

**Authors:** Nathan J. Cheetham, Rose Penfold, Valentina Giunchiglia, Vicky Bowyer, Carole H. Sudre, Liane S. Canas, Jie Deng, Benjamin Murray, Eric Kerfoot, Michela Antonelli, Khaled Rjoob, Erika Molteni, Marc F. Österdahl, Nicholas R. Harvey, William R. Trender, Michael H. Malim, Katie J. Doores, Peter J. Hellyer, Marc Modat, Alexander Hammers, Sebastien Ourselin, Emma L. Duncan, Adam Hampshire, Claire J. Steves

## Abstract

**Background:** Cognitive impairment has been reported after many types of infection, including SARS-CoV-2. Whether deficits following SARS-CoV-2 improve over time is unclear. Studies to date have focused on hospitalised individuals with up to a year follow-up. The presence, magnitude, persistence and correlations of effects in community-based cases remain relatively unexplored.

**Methods:** Cognitive performance (working memory, attention, reasoning, motor control) was assessed in participants of a voluntary biobank in July, 2021 (Round 1), and April, 2022 (Round 2). Participants, drawn from the COVID Symptom Study smartphone app, comprised individuals with and without SARS-CoV-2 infection and varying symptom duration. Effects of COVID-19 exposures on cognitive accuracy and reaction time scores were estimated using multivariable ordinary least squares linear regression models weighted for inverse probability of participation, adjusting for potential confounders and mediators. The role of ongoing symptoms after COVID-19 infection was examined stratifying for self-perceived recovery. Longitudinal analysis assessed change in cognitive performance between rounds.

**Findings:** 3335 individuals completed Round 1, of whom 1768 also completed Round 2. At Round 1, individuals with previous positive SARS-CoV-2 tests had lower cognitive accuracy (N = 1737, β = −0.14 standard deviations, SDs) than negative controls. Deficits were largest for positive individuals with ≥ 12 weeks of symptoms (N = 495, β = −0.22 SDs). Effects were comparable to hospital presentation during illness (N = 281, β = −0.31 SDs), and 10 years age difference (60-70 years vs. 50-60 years, β = −0.21 SDs) in the whole study population. Stratification by self-reported recovery revealed that deficits were only detectable in SARS-CoV-2 positive individuals who did not feel recovered from COVID-19, whereas individuals who reported full recovery showed no deficits. Longitudinal analysis showed no evidence of cognitive change over time, suggesting that cognitive deficits for affected individuals persisted at almost 2 years since initial infection.

**Interpretation:** Cognitive deficits following SARS-CoV-2 infection were detectable nearly two years post infection, and largest for individuals with longer symptom durations, ongoing symptoms, and/or more severe infection. However, no such deficits were detected in individuals who reported full recovery from COVID-19. Further work is needed to monitor and develop understanding of recovery mechanisms for those with ongoing symptoms.

**Funding:** Chronic Disease Research Foundation, Wellcome Trust, National Institute for Health and Care Research, Medical Research Council, British Heart Foundation, Alzheimer’s Society, European Union, COVID-19 Driver Relief Fund, French National Research Agency.

**Research in context:** *Evidence before this study:* Abstracts were screened from a PubMed search query (COVID-19) AND (long COVID) AND (cognitive impairment), which returned 409 results between 2020 and January 20, 2023. Multiple systematic reviews and meta-analyses reported consistent observation of cognitive deficits following SARS-CoV-2 infection. Most studies of cognitive impairment have used small samples of less than 200 participants (including any controls), hospitalised cohorts, and measured cognitive impairment through self-report or dichotomised quantitative scales. Only one study was found with a sample size of more than 1,000 individuals, included cases and controls across both community and hospital settings, and used objective cognitive testing that allowed quantitative estimation of the scale of any cognitive impairment. Previous studies have also been limited insofar as focusing on earlier infections in the first year of the COVID-19 pandemic, prior to introduction of vaccination and emerging variants. Studies focusing on longitudinal follow-up for those hospitalised with COVID-19 or with long COVID have found low rates of full recovery from long-term symptoms at up to one year since infection, including cognitive impairment.

*Added value of this study:* We report quantitatively on cognitive impairment following SARS-CoV-2 infection, from a large dataset of 4,000 individuals with and without test-confirmed SARS-CoV-2 infection and a range of associated symptom durations, with mostly community-based cases. Importantly, we undertook two rounds of cognitive testing allowing longitudinal tracking of cognitive performance. Our longitudinal methods allowed us to report on deficits up to two years since infection, and following infections with SARS-CoV-2 variants that have emerged over 2021 and 2022, not previously studied in the context of COVID-19 and cognition.

*Implications of all the available evidence:* This study adds to existing evidence of cognitive deficits following SARS-CoV-2 infection, but finds important exceptions. At initial testing in mid-2021, cognitive deficits are not found for individuals who self-report as feeling recovered from COVID-19, even for those with longest symptom duration. In follow-up testing in mid-2022, we find that deficits appear persistent for those with earlier infections and ongoing symptoms, consistent with previous smaller studies. More research is required to monitor those experiencing persistent cognitive impairment and understand the mechanisms underlying recovery.

## Introduction

Persistent cognitive impairment and cognitive deficits after SARS-CoV-2 infection in comparison to individuals without infection have been reported from both subjective self-reported survey and objective assessments of cognitive functioning [1–3]. Effects are similar to other infections [4,5]. Studies using objective assessment to quantify the magnitude of cognitive deficits found deficits increased with severity of illness during the acute phase, with deficits among individuals requiring respiratory support or mechanical ventilation similar in magnitude to ageing 20 years from 50 to 70 years [6,7]. A UK Biobank study comparing magnetic resonance images and objective cognitive tests recorded before and after SARS-CoV-2 infection revealed structural brain changes and longitudinal decline in cognitive performance [8], while markers of brain injury have been found in hospitalised COVID-19 patients [9].

In addition to cognitive impairment in hospitalised individuals, cognitive impairment has also been reported in individuals with long-term and/or ongoing symptoms following SARS-CoV-2 infection, referred to most commonly as long COVID (clinical definitions and terms vary but generally refer to symptoms associated with SARS-CoV-2 infection which persist for more than 4 or 12 weeks since infection [10,11]) [1,3]. While 17% of a UK cohort of hospitalised individuals met criteria of cognitive impairment from objective assessment at 6 months since hospital discharge [12], the UK Office for National Statistical estimated in January, 2023 that 1.0 million (52%) and 771,000 (39%) of 2.0 million individuals in the UK with self-reported long COVID (symptoms persisting for more than four weeks since infection) were experiencing difficulty concentrating and memory loss or confusion respectively [13]. Ongoing symptoms experienced by individuals with long COVID are associated with difficulties in daily functioning, reduced ability to work, lower mental health and wellbeing, and lower self-reported quality-of-life [14–18]. As well as effects on current functioning, SARS-CoV-2 infection may accelerate cognitive decline with age. A large-scale international study using electronic healthcare records to identify cases found increased risk of cognitive deficit and dementia persisting for at least two years after recorded COVID-19 diagnosis in comparison to other respiratory infections [19].

Previous studies using objective cognitive assessments have mostly examined small, hospitalised cohorts, used dichotomised classifications rather than quantitative scales of cognitive impairment, focussed on individuals infected in the first year of the pandemic (i.e., before vaccination), with generally short follow-up (typically 6-12 months since infection) [1–3]. While studies have shown neurological deficits following SARS-CoV-2 infection, including in cohorts with long COVID, to our knowledge no studies have analysed a cohort with a range of symptom durations for both SARS-CoV-2 positive cases and negative controls, to examine the effects of both SARS-CoV-2 infection and symptom duration. Furthermore, few studies have looked longitudinally at cognitive trajectories of individuals and whether recovery relates to cognitive performance.

In this study, we used a validated cognitive assessment tool, with prospective self-report symptom assessment, and retrospective reflective survey data from a large UK voluntary cohort, the COVID Symptom Study Biobank, to address the following questions: 1) Is COVID-19 associated with cognitive performance? 2) Do symptom duration and ongoing symptoms affect any observed associations between COVID-19 and cognitive performance? 3) Do any associations between COVID-19 and cognitive performance change over time? We hypothesise that longer COVID-19 symptom duration has larger detriments to cognitive performance, while individuals reporting recovery from COVID-19 will show reduced cognitive deficits in comparison to those with ongoing symptoms.

## Methods

### COVID Symptom Study Biobank cohort

Study participants were volunteers from the COVID Symptom Study Biobank (CSSB), approved by Yorkshire & Humber NHS Research Ethics Committee Ref: 20/YH/0298. Individuals were recruited to the CSSB via the COVID Symptom Study (CSS, later renamed ZOE Health Study) launched in the UK on March 24, 2020, approved by the King’s College London Ethics Committee LRS-19/20-18210. All data were collected with informed consent obtained online. Via the CSS app, participants self-report demographic information, symptoms potentially suggestive of COVID-19, any SARS-CoV-2 testing and results, and any vaccinations. CSS participants from across the UK were invited to join the CSSB by email in October to November, 2020 and May, 2021.

CSSB invitation targeted five groups of regular CSS users with different SARS-CoV-2 infection statuses and associated symptom (illness) durations at the time of invitation. Case group 1 comprised individuals with positive SARS-CoV-2 test but no associated symptoms (asymptomatic COVID). Case group 2 comprised individuals with positive SARS-CoV-2 test and between one and 13 days of associated symptoms (short COVID). Case group 3 comprised individuals with positive SARS-CoV-2 test and at least 28 days of associated symptoms (long COVID). Control group 1 comprised individuals with negative SARS-CoV-2 test and at least 28 days of symptoms at the time of the test (long non-COVID). Control group 2 comprised individuals with negative SARS-CoV-2 test associated with one to maximum three consecutive days of illness at the time of the test, with low symptom burden (three symptoms or fewer; healthy non-COVID). Invited individuals were matched by Euclidian distance for age, sex and body mass index (BMI) across groups. Due to this targeted approach designed to give five equally sized groups, cohort composition is not representative of population prevalence of COVID-19 and long COVID. After consent, all participants were invited to give blood samples that were tested for anti-Nucleocapsid and anti-Spike SARS-CoV-2 antibodies.

8,357 CSSB participants were invited to participate in a first round cognitive testing in July, 2021. Participants (whether participating in the first round or not) were also invited to a second testing round in April, 2022.

### Data collection

#### Cognitive assessment

Cognitive assessment was performed using the “Cognitron” online platform https://www.cognitron.co.uk/. In addition to studies of both hospitalised and community cases of COVID-19 [6,7], cognitive batteries using the same platform have previously been shown to be sensitive to cognitive impairment in early Alzheimer’s disease, cognitive decline in individuals with mild behavioural impairment (an at-risk state for dementia), cognitive function in older adults with high vs. low autistic traits, and “brain training” task repetition [20–24]. Cognitive impairments following traumatic brain injury measured using the platform have also been found to correlate with corresponding brain networks measured by MRI [25].

In both rounds of assessment, participants undertook 12 cognitive tasks assessing different cognitive domains, including working memory, attention, reasoning, and motor control. Accuracy, average within-task reaction time, and variation in within-task reaction time metrics were extracted for each task and participant. Cognitive domain tested, performance metrics and transformation methods for each task are given in Table S1.

Concurrently with cognitive testing through the same online platform, participants completed the following assessments: Patient Health Questionnaire-2 (PHQ-2) and Generalised Anxiety Disorder-7 (GAD-7) assessments of depression and anxiety symptoms [26,27], Chalder Fatigue Scale (CFS) [28,29], and Work and Social Adjustment Scale (WSAS) assessing functional impairment [30]. a The PHQ-4 measure of psychological distress was generated from the PHQ-2 and GAD-7 [31]. During second testing round only, individuals were also asked to report their highest level of educational attainment, as this is known to affect task performance.

#### Other data sources

The following variables were derived from self-reported data at registration with the CSS app: age, biological sex, ethnicity, physical health conditions (asthma, cancer, diabetes, heart disease, kidney disease, lung disease), weight and height (from which BMI was derived), frailty (from the PRISMA-7 scale [32]), local area deprivation (Index of Multiple Deprivation, IMD [33]), and UK geographic region (from residential address data). Age, ethnicity, height, weight, and residential address were re-collected at time of consent to join CSSB and superseded data collected in the CSS app. SARS-CoV-2 testing data, symptoms data and whether individuals presented to hospital over the course of any illness were collected from the CSS app using ExeTera software (extraction date: 30 May, 2022) [34]. Number of mental health conditions was measured from self-reported diagnoses of 16 conditions collected in a February, 2021 CSS questionnaire. SARS-CoV-2 antibody testing results were generated from blood samples collected after consent at King’s College London using previously reported methods [35]. Self-perceived recovery from COVID-19 was collected in May-June, 2021 shortly before cognitive testing Round 1, as part of the CSSB “Effects of the Coronavirus Disease (COVID-19) pandemic on life in the UK” questionnaire.

A “COVID-19 group” exposure variable was derived as a composite combining SARS-CoV-2 test result (infection status) and duration of symptoms associated with the test (symptoms starting within a 14 day window either side of the positive or negative antigen test or more than 14 days before an antibody test), adapting methodology from a previous report (criteria provided in supplementary information section S2 and visualised in Figure S1) [36]. COVID-19 groups at both Round 1 and 2 of cognitive assessment were derived based on SARS-CoV-2 test and symptom data up to date of invitation to Round 1 and Round 2 respectively, allowing for changes in COVID-19 group between rounds due to new infection and/or symptoms. Symptom duration estimates were categorised based on COVID-19 clinical case definitions recommended in NICE guidelines on managing the long-term effects of COVID-19 [10]: asymptomatic, symptoms up to 4 weeks (analogous to NICE “acute COVID-19”), symptoms between 4 and 12 weeks (analogous to NICE “ongoing symptomatic COVID-19”), and symptoms for 12 weeks or more (analogous to NICE “post-COVID-19 syndrome”). Equivalent symptom duration groupings associated with both negative and positive tests were used to give a total of 8 categories for COVID-19 group.

#### Inclusion criteria

For all analyses, inclusion criteria were complete age, sex, ethnicity and area of residence data, and sufficient self-reported SARS-CoV-2 test results and symptom assessments logged on the CSS app at the time of invitation to Round 1 cognitive assessment to derive and assign a “COVID-19 group” described above. Full completion of all cognitive tasks was further required for generation of composite scores used as primary outcomes.

### Statistical methods

#### Generation of inverse participation weights

Preliminary analysis of cognitive testing participation rates showed variation with our primary exposures of interest (SARS-CoV-2 infection status and associated symptom duration). Knowing this may act as a potential source of response and collider bias (as demonstrated in other COVID-19 research [37]), logistic regression models were run to predict participation in cognitive assessments and generate weights of inverse probability of participation. Weights generated from these models were included in subsequent analyses in attempt to account for bias. Further details of weight generation are given in supplementary information section S3.

#### Cognitive assessment data processing

For the small number of individuals who completed a task more than once within the same testing round (due to multiple attempts to complete the cognitive assessment), task metrics were taken for the earliest attempt to prevent learning curve effects. Reaction time metrics more than three standard deviations from the mean were winsorised to reduce effects of outliers as done previously [6]. Transformation methods were applied to each task metric (square, cube, square root or logarithmic), to minimise skewness of distributions (Table S1). Following transformation, metrics for each task were standardised into z-scores representing number of standard deviations from the mean.

#### Principal component analysis

Principal component analysis (PCA) was used to generate composite scores for accuracy, average reaction time, and reaction time variation across all cognitive tasks. PCA was performed separately on Round 1 and Round 2 datasets, with the number of components selected using a previously reported method [38]. Principal component composite scores were converted to standardised z-scores for use in subsequent analyses.

#### Proposed causal pathways: Directed acyclic graphs

Using a causal inference approach to estimate the individual effects of various exposure variables on both participation in cognitive assessment and cognitive performance, directed acyclic graphs (DAGs) describing proposed relationships between observed variables were constructed using dagitty software http://www.dagitty.net/dags.html (Figure 1, full code given in supplementary information section S5) [39]. To estimate the total causal effect of each exposure listed on the outcomes of interest (i.e., participation and cognitive performance), separate models were run for each exposure with adjustment variable sets suggested from the proposed DAGs (adjustment sets tabulated in Table S2). In this way, the so-called “Table 2 fallacy” (misinterpretation of effect estimates of adjustment variables often presented in multivariable model results as if they were the exposure of interest – in which case they may require a different adjustment set) can be avoided [40]. Relationships between variables shown in DAGs were informed by previously observed associations wherever possible. For analyses of individuals who participated in Round 1 cognitive testing, local area deprivation and UK geographic region were used as proxy variables for educational attainment level, which was only collected in Round 2 of assessment. For analyses of individuals that completed Round 2 or both Round 1 and 2, educational attainment level collected at Round 2 was used with the assumption that educational attainment level was constant.

**Figure 1.**
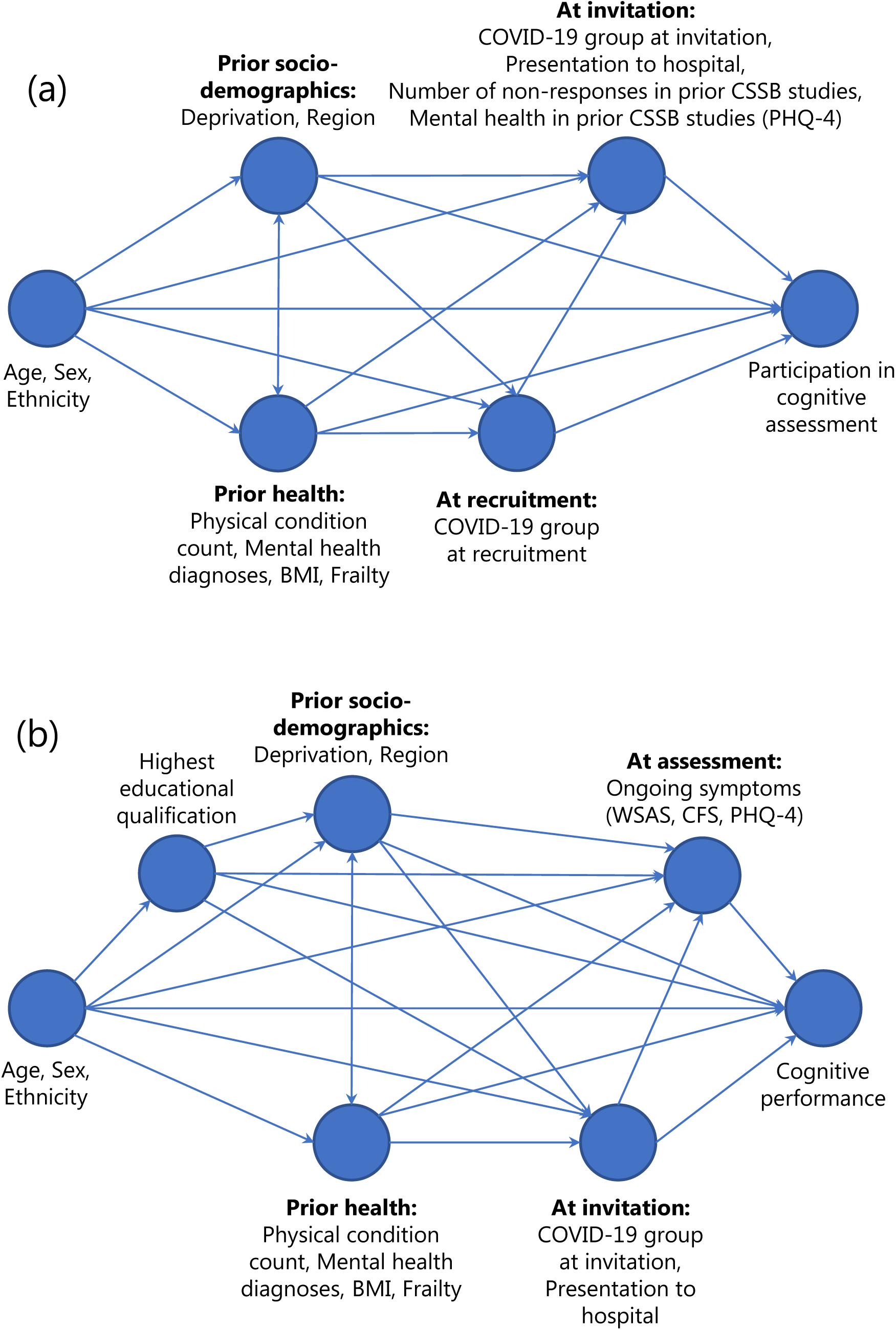
Directed acyclic graph describing hypothesised causal pathways. Proposed directed acyclic graphs (DAGs) used to generate minimal adjustment variable sets for estimation of the total causal effect of variables on outcomes of participation in cognitive assessment (a) and cognitive performance (b). DAGs are structured approximately in order of data generation from left to right.

#### Regression models

The total effects of exposure variables on participation in cognitive testing were estimated using ordinary logistic regression models containing the outcome and exposure variable of interest, in addition to the appropriate adjustment set (Figure 1).

The total effects of exposure variables on cognitive performance metrics (accuracy, reaction time and variation in reaction) were estimated using ordinary least squares linear regression models containing the outcome and exposure variable of interest, in addition to the appropriate adjustment set (Figure 1). Separate models were run for each combination of outcome and exposure variable. To estimate the direct effect of COVID-19 group on change in cognitive performance from Round 1 to Round 2, scores from Round 1 for the equivalent Round 2 outcome metric were included as additional variables in models. This approach was used instead of using a “change score” as an outcome, due to lack of meaningful interpretation [41]. For all models with the same outcome, p-values were adjusted for multiple testing using the Benjamini/Hochberg method [42].

#### Software

Analyses were performed using python v3.8.8 and packages: numpy v1.20.1, pandas v1.2.4, statsmodels v0.12.2, scipy v1.6.2, scikit-learn v0.24.1, matplotlib v3.3.4, seaborn v0.11.1, factor_analyzer v0.4.0.

## Results

### Sample characteristics

Of 8,357 individuals invited to Round 1 and/or Round 2 of cognitive assessment, 7,588 Round 1 invitees and 7,198 Round 2 invitees met inclusion criteria for analysis (Figure S2). Of these, 3,335 completed all cognitive assessment tasks in Round 1, 2,435 in Round 2, and 1,768 in both rounds.

Participants in cognitive assessments were skewed towards middle age groups (Round 1: median = 57 years [IQR = 50-64], Round 2: median = 58 years [IQR = 51-64]), female sex (Round 1: 81%,

Round 2: 82%), white ethnicity (Round 1: 96%, Round 2: 97%), living in lower deprivation neighbourhoods (IMD Quintile 5, least deprived 20%, Round 1: 35%, Round 2: 34%), reflecting CSSB cohort composition (Table 1). Half of Round 1 participants (52%) had self-reported one or more positive SARS-CoV-2 tests at time of Round 1 invitation (July 12, 2021), increasing to two-thirds (67%) in Round 2 participants at time of Round 2 invitation, (April 28, 2022). The largest COVID-19 groups were individuals with a positive test and symptoms of up to two weeks, Round 1: 18%, Round 2: 27% (meeting NICE “acute-COVID-19” criteria [10]) and individuals with a positive test and symptoms lasting 12 or more weeks, Round 1: 15%, Round 2: 16% (meeting NICE “post-COVID-19 syndrome” criteria). The number of individuals in the “healthy control” COVID-19 group, with only negative test(s) and no associated symptoms, decreased between Rounds 1 and 2 because of ongoing SARS-CoV-2 infection in the general population (Round 1: 19%, Round 2: 9%). Sample characteristics stratified by COVID-19 group are given in Table S3. In line with CSSB recruitment based on symptom duration status during UK wild-type or alpha-variant waves in 2020, almost all individuals with positive tests and associated symptoms of 12 or more weeks were infected in 2020 (Round 1: 98%, Round 2: 91%).

**Table 1.** Sample characteristics. Characteristics of participants in Round 1 and/or Round 2 of cognitive assessment. Counts and proportions are presented apart from weeks between cognitive assessment and symptom start/test date, where median and interquartile range is given. For variables relating to COVID-19 history, marked with *, counts are given at the time of invitation to the relevant round (counts at invitation to Round 1 for column detailing participants in both rounds). Education level was collected as part of Round 2 of cognitive assessment. Other variables were collected or derived from data collected at recruitment or prior to analyses.

| Variable | Category | Invited to Round 1 & meeting inclusion criteria | Round 1 | Round 2 | Full completion of both Round 1 and 2 |
| --- | --- | --- | --- | --- | --- |
| **TOTAL COUNT** |  | 7588 | 3335 | 2435 | 1768 |
| Age group (years) | 18-30 | 166 (2.2%) | 43 (1.3%) | 23 (0.9%) | 15 (0.8%) |
|  | 30-40 | 695 (9.2%) | 214 (6.4%) | 128 (5.3%) | 80 (4.5%) |
|  | 40-50 | 1490 (19.6%) | 539 (16.2%) | 379 (15.6%) | 258 (14.6%) |
|  | 50-60 (reference) | 2572 (33.9%) | 1183 (35.5%) | 860 (35.3%) | 622 (35.2%) |
|  | 60-70 | 2059 (27.1%) | 1042 (31.2%) | 804 (33.0%) | 613 (34.7%) |
|  | 70-80 | 570 (7.5%) | 293 (8.8%) | 223 (9.2%) | 168 (9.5%) |
|  | ≥ 80 | 36 (0.5%) | 21 (0.6%) | 18 (0.7%) | 12 (0.7%) |
| Sex | Female (reference) | 6032 (79.5%) | 2697 (80.9%) | 2001 (82.2%) | 1445 (81.7%) |
|  | Male | 1556 (20.5%) | 638 (19.1%) | 434 (17.8%) | 323 (18.3%) |
| Ethnicity | Asian/Asian British | 61 (0.8%) | 25 (0.7%) | 14 (0.6%) | 11 (0.6%) |
|  | Black/Black British | 27 (0.4%) | 12 (0.4%) | 5 (0.2%) | < 5 |
|  | Mixed/Multiple | 98 (1.3%) | 41 (1.2%) | 25 (1.0%) | 19 (1.1%) |
|  | Other | 111 (1.5%) | 44 (1.3%) | 26 (1.1%) | 18 (1.0%) |
|  | White (reference) | 7291 (96.1%) | 3213 (96.3%) | 2365 (97.1%) | 1717 (97.1%) |
| Highest educational attainment level | Data not available | 4422 (58.3%) | 1194 (35.8%) | 0 (0.0%) | 0 (0.0%) |
|  | Postgraduate degree or higher | 959 (30.3%) | 658 (30.7%) | 759 (31.2%) | 557 (31.5%) |
|  | Undergraduate degree (reference) | 1169 (36.9%) | 794 (37.1%) | 900 (37.0%) | 653 (36.9%) |
|  | Less than undergraduate degree | 994 (31.4%) | 661 (30.9%) | 740 (30.4%) | 534 (30.2%) |
|  | Other/Prefer not to say | 44 (1.4%) | 28 (1.3%) | 36 (1.5%) | 24 (1.4%) |
| Local area deprivation (IMD) | Quintile 1 (most 20% deprived areas) | 470 (6.2%) | 188 (5.6%) | 138 (5.7%) | 97 (5.5%) |
|  | Quintile 2 | 1014 (13.4%) | 413 (12.4%) | 335 (13.8%) | 235 (13.3%) |
|  | Quintile 3 (reference) | 1517 (20.0%) | 682 (20.4%) | 486 (20.0%) | 359 (20.3%) |
|  | Quintile 4 | 2010 (26.5%) | 883 (26.5%) | 640 (26.3%) | 469 (26.5%) |
|  | Quintile 5 (least 20% deprived areas) | 2577 (34.0%) | 1169 (35.1%) | 836 (34.3%) | 608 (34.4%) |
| SARS-CoV-2 test result* | Negative (reference) | 3453 (45.5%) | 1598 (47.9%) | 1150 (47.5%) | 854 (48.3%) |
|  | Positive | 4135 (54.5%) | 1737 (52.1%) | 1273 (52.5%) | 914 (51.7%) |
| COVID-19 group* | SARS-CoV-2 Negative, Asymptomatic (reference) | 1139 (15.0%) | 641 (19.2%) | 482 (19.8%) | 388 (21.9%) |
|  | SARS-CoV-2 Negative, Symptom duration < 4 weeks | 919 (12.1%) | 379 (11.4%) | 271 (11.1%) | 190 (10.7%) |
|  | SARS-CoV-2 Negative, Symptom duration 4-12 weeks | 977 (12.9%) | 372 (11.2%) | 259 (10.6%) | 175 (9.9%) |
|  | SARS-CoV-2 Negative, Symptom duration ≥ 12 weeks | 418 (5.5%) | 206 (6.2%) | 136 (5.6%) | 101 (5.7%) |
|  | SARS-CoV-2 Positive, Asymptomatic | 706 (9.3%) | 256 (7.7%) | 176 (7.2%) | 122 (6.9%) |
|  | SARS-CoV-2 Positive, Symptom duration < 4 weeks | 1601 (21.1%) | 589 (17.7%) | 402 (16.5%) | 279 (15.8%) |
|  | SARS-CoV-2 Positive, Symptom duration 4-12 weeks | 985 (13.0%) | 397 (11.9%) | 298 (12.2%) | 209 (11.8%) |
|  | SARS-CoV-2 Positive, Symptom duration ≥ 12 weeks | 843 (11.1%) | 495 (14.8%) | 370 (15.2%) | 304 (17.2%) |
| Presented to hospital during symptomatic period* | No (reference) | 7003 (92.3%) | 3054 (91.6%) | 2172 (90.7%) | 1594 (90.2%) |
|  | Yes | 585 (7.7%) | 281 (8.4%) | 223 (9.3%) | 174 (9.8%) |
| Weeks between cognitive assessment and symptom start/test date* |  | 53.0 (31.0, 67.0) | 42.0 (29.0, 62.0) | 78.0 (38.0, 103.0) | 77.0 (40.0, 101.0) |
| Symptom start date/Test date* | Q1 Jan-Mar, 2020 | 933 (12.3%) | 385 (11.5%) | 263 (10.8%) | 187 (10.6%) |
|  | Q2 Apr-Jun, 2020 | 2497 (32.9%) | 879 (26.4%) | 511 (21.0%) | 332 (18.8%) |
|  | Q3 Jul-Sep, 2020 | 1065 (14.0%) | 469 (14.1%) | 232 (9.5%) | 179 (10.1%) |
|  | Q4 Oct-Dec, 2020 | 1746 (23.0%) | 830 (24.9%) | 500 (20.5%) | 386 (21.8%) |
|  | Q1 Jan-Mar, 2021 | 603 (7.9%) | 319 (9.6%) | 129 (5.3%) | 102 (5.8%) |
|  | Q2 Apr-Jun, 2021 | 712 (9.4%) | 433 (13.0%) | 155 (6.4%) | 126 (7.1%) |
|  | Q3 Jul-Sep, 2021 | 32 (0.4%) | 20 (0.6%) | 78 (3.2%) | 57 (3.2%) |
|  | Q4 Oct-Dec, 2021 | 0 | 0 | 196 (8.0%) | 141 (8.0%) |
|  | Q1 Jan-Mar, 2022 | 0 | 0 | 305 (12.5%) | 212 (12.0%) |
|  | Q2 Apr-Jun, 2022 | 0 | 0 | 66 (2.7%) | 46 (2.6%) |

### Participation analysis

Logistic regression analysis of factors associated with participation in cognitive assessments revealed multiple associations, with individuals with a higher number of prior non-responses to other CSSB studies, more recently recruited individuals, older age groups, and female sex individuals more likely to participate (Figure S3, Figure S4, Table S4). Predictive performance of models used to generate weights of inverse probability of participation for subsequent analyses was moderate for Round 1 participation (AUC-ROC = 0.72), and good for Round 2 participation (AUC-ROC = 0.82) and participation in both Round 1 and 2 (AUC-ROC = 0.87) (Table S5).

### Principal component analysis of cognitive performance

Separate principal component analyses (PCA) of accuracy, average reaction time, and intra-task reaction time variation metrics from Round 1 and 2 of cognitive assessment produced three, two, and three components with an eigenvalue greater than one respectively (Table S6 and Table S7). Very similar loadings and variance explained were found for the first and second principal components for each performance metric across both rounds of testing. As a result, loadings from Round 1 PCA were used to generate composite scores for first and second principal components for both Round 1 and 2, for each of accuracy, average reaction time, and reaction time variation. The first principal component from PCA of task accuracy scores represented higher accuracy across all tasks (26% of variance). The first component from average task reaction time PCA represented larger average reaction time (44% of variance), and first component of variation in reaction time PCA represented larger reaction time variation (25% of variance). First component scores were used as the primary outcomes of interest in subsequent analyses.

### Cross-sectional cognitive performance

At Round 1 of cognitive testing, lower composite cognitive accuracy scores, or “cognitive deficits”, were found for the SARS-CoV-2 positive COVID-19 group with ≥ 12 weeks symptom duration (β = −0.22 standard deviations from mean [SDs], 95% CI: −0.35, −0.09, adjusted p = 0.005), relative to the SARS-CoV-2 negative COVID-19 group with no symptoms (Figure 2). The scale of deficit was similar to the effect size of presentation to hospital during illness (β = −0.31 SDs, 95% CI: −0.44, −0.18, adjusted p < 0.0001), or of age group differences of 10 years within the same sample, e.g., age 60-70 years vs. 50-60 years (β = −0.21 SDs, 95% CI: −0.30, −0.13, adjusted p < 0.0001), or age 40-50 years vs. 50-60 years (β = +0.24 SDs, 95% CI: 0.15, 0.34, adjusted p < 0.0001) (Figure S5). While controlling for symptom duration, an overall deficit was observed for individuals with positive SARS-CoV-2 tests compared to individuals testing negative (β = −0.14 SDs, 95% CI: −0.21, −0.07, adjusted p = 0.0003). Cognitive deficits in SARS-CoV-2 positive COVID-19 groups remained present in the subset of individuals who completed both Round 1 and 2 of assessment, for whom educational attainment data was available and included as an additional adjustment variable (Figure S6). There was no evidence of cognitive deficits in individuals in symptomatic SARS-CoV-2 negative COVID-19 groups.

**Figure 2.**
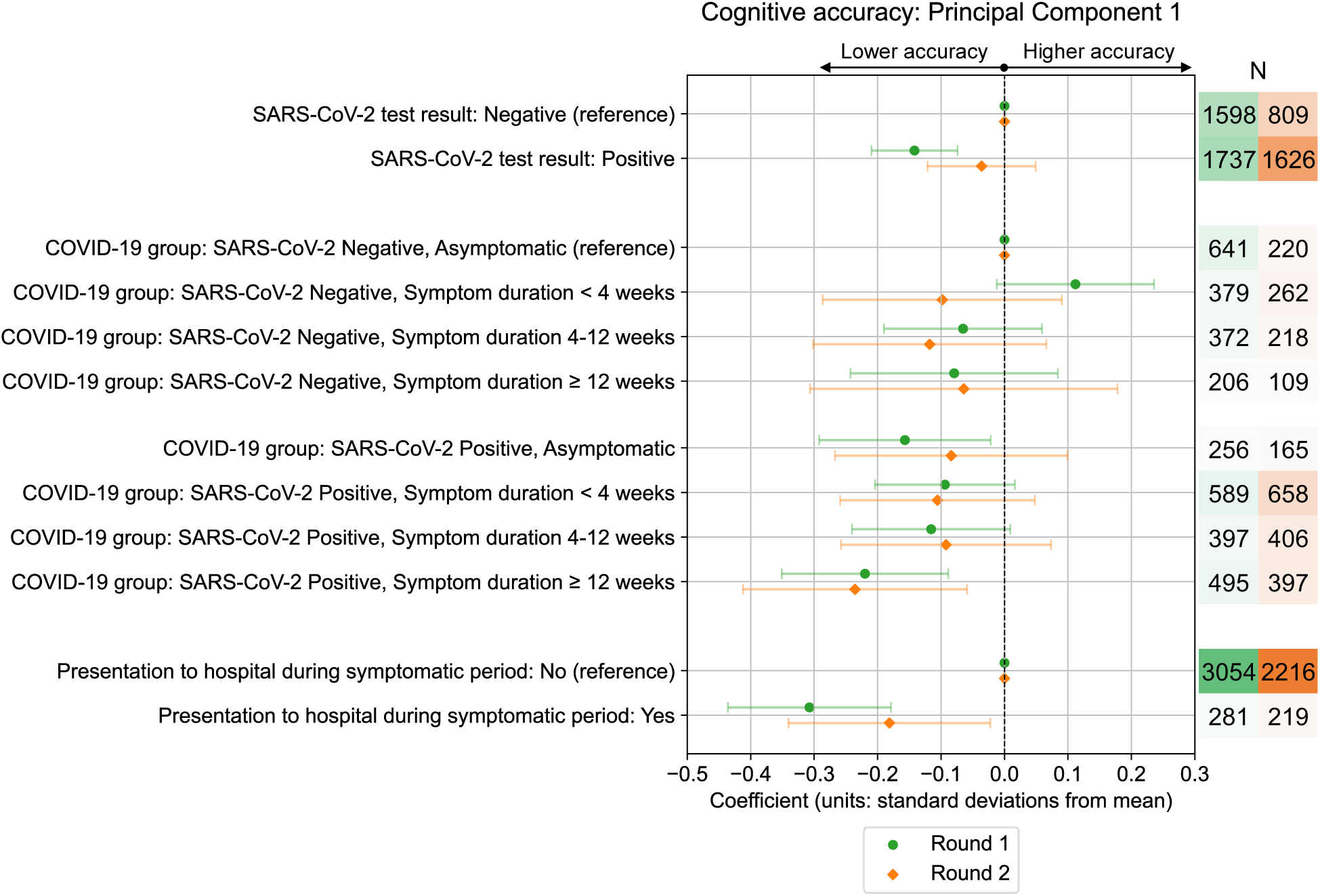
Association between COVID-19 exposures and cognitive accuracy scores. Standardised coefficients (number of standard deviations from mean) with 95% confidence intervals from multivariable ordinary least squares linear regression models testing association between COVID-19 related exposures and cognitive accuracy PCA 1^st^ component standardised scores. Results are from Round 1 and Round 2 of cognitive testing, among all individuals who completed either round of testing. Results for each exposure variable presented originate from separate models that use distinct adjustment variable sets determined from the proposed DAG for cognitive performance (Test result – Age, BMI, Deprivation, Education [Round 2 model only], Ethnicity, Frailty, Mental health condition count, Physical health condition count, Presentation to hospital, Region, Sex, Symptom duration; COVID-19 group – Age, BMI, Deprivation, Education [Round 2 model only], Ethnicity, Frailty, Mental health condition count, Physical health condition count, Presentation to hospital, Region, Sex; Presentation to hospital – Age, BMI, Deprivation, Education [Round 2 model only], Ethnicity, Frailty, Mental health condition count, Physical health condition count, Presentation to hospital, Region, Sex, Symptom duration).

At Round 2 of testing, lower cognitive accuracy scores were again observed for the SARS-CoV-2 positive COVID-19 group with ≥ 12 weeks symptom duration, and individuals who presented to hospital during illness. However, effect sizes and/or strength of associations (wider confidence intervals and larger p-values) with SARS-CoV-2 infection were generally reduced in comparison to Round 1.

Analysing associations with accuracy in individual tasks, deficits due to COVID-19 exposures were largest in tasks focusing on working memory (Figure S7). Further multivariable models found several other exposures were associated with lower cognitive accuracy for both principal components composites and individual task scores, namely older age, lower educational attainment level, living in certain UK regions, and areas of higher deprivation (used as socio-economic indicators in models of all Round 1 participants for whom educational data was incomplete), BMI in the underweight, overweight or obese range, having one or more physical health conditions (from asthma, cancer, diabetes, heart disease, kidney disease, lung disease), and above threshold scores indicative of poorer mental health, high fatigue levels, and functional impairment in assessments reported contemporaneously with cognitive assessments (Figure S5 and Figure S8). Multivariable models testing associations with within-task average reaction time and reaction time variation found higher within-task variation in reaction time for SARS-CoV-2 positive individuals with ≥ 12 weeks symptom duration in Round 1 (Figure S9), but no evidence of differences in average reaction time among SARS-CoV-2 positive groups (Figure S10).

### Role of ongoing symptoms

To test the role of ongoing symptoms in the observed cognitive deficits, further models testing associations with Round 1 cognitive accuracy were run. Firstly, the role of self-perceived recovery from COVID-19 was tested by stratifying SARS-CoV-2 positive individuals by their response to the question “Thinking about the last or only episode of COVID-19 you have had, have you now recovered and are back to normal?” in a separate CSSB survey undertaken shortly before Round 1, completed by 84% (N = 1455/1737) of SARS-CoV-2 positive individuals at Round 1 (Figure 3).

**Figure 3.**
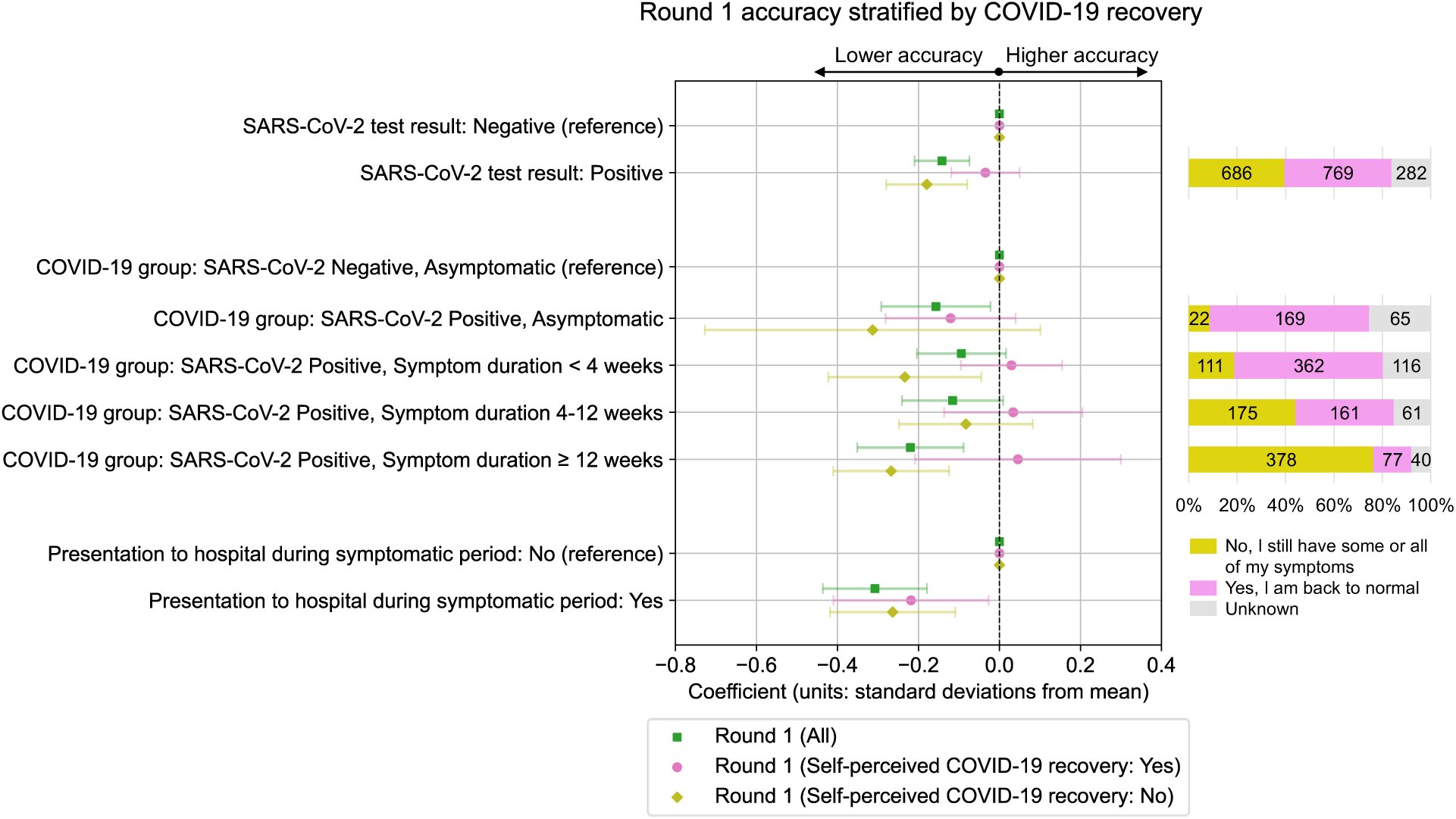
Association between COVID-19 exposures and Round 1 cognitive accuracy scores, stratified by self-perceived recovery from COVID-19. Standardised coefficients (number of standard deviations from mean) with 95% confidence intervals from multivariable ordinary least squares linear regression models testing association between COVID-19 related exposures and cognitive accuracy PCA 1^st^ component standardised scores. Results are from all individuals who completed Round 1 of cognitive testing, stratified by response to the question “Thinking about the last or only episode of COVID-19 you have had, have you now recovered and are back to normal?”, with cross-tabulations presented for SARS-CoV-2 positive individuals. Results for each exposure variable presented originate from separate models that use distinct adjustment variable sets determined from the proposed DAG for cognitive performance (Test result – Age, BMI, Deprivation, Ethnicity, Frailty, Mental health condition count, Physical health condition count, Presentation to hospital, Region, Sex, Symptom duration; COVID-19 group – Age, BMI, Deprivation, Ethnicity, Frailty, Mental health condition count, Physical health condition count, Presentation to hospital, Region, Sex; Presentation to hospital – Age, BMI, Deprivation, Ethnicity, Frailty, Mental health condition count, Physical health condition count, Presentation to hospital, Region, Sex, Symptom duration).

After stratification, cognitive deficits were not observed among SARS-CoV-2 positive individuals (N = 769) who responded “Yes, I am back to normal”, both for individuals with ≥ 12 weeks symptoms (β = +0.05 SDs, 95% CI: −0.21, 0.3, adjusted p = 0.86) and overall while controlling for symptom duration (β = −0.03 SDs, 95% CI: −0.12, 0.05, adjusted p = 0.72). Conversely, overall deficits in SARS-CoV-2 positive were increased compared to the unstratified sample for the 686 individuals who responded “No, I still have some or all of my symptoms” (β = −0.18 SDs, 95% CI: −0.28, −0.08, adjusted p = 0.003). However, self-perceived COVID-19 recovery among SARS-CoV-2 positive individuals was highly correlated with symptom duration (Figure S12, Pearson correlation coefficient for weeks of symptoms vs. proportion recovered, r = −0.77, p = 2 × 10^−11^).

Individuals who presented to hospital during illness showed a similar scale of cognitive deficits independent of self-perceived recovery from COVID-19. A small proportion of individuals (22/256, 9%) who did not report any symptoms prospectively via the CSS app at the time of their SARS-CoV-2 infection and so were considered as asymptomatic reported as “still having some or all of their symptoms” when asked retrospectively about their recovery from COVID-19 infection.

Further analyses used mental health, fatigue, and functional impairment assessments collected contemporaneously with cognitive assessment to test mediation of cognitive deficits by specific symptom types. Associations between COVID-19 group and cognitive accuracy in mediation models showed reduced effect sizes and 95% confidence levels which crossed β = 0 for some groups in both Round 1 and Round 2 (Figure S11). Results suggest partial mediation by these symptom types on the effect of COVID-19 group on cognitive accuracy, with mediation effects larger for longer symptom duration groups.

### Longitudinal change in cognitive accuracy between rounds

Finally, to estimate the effect of COVID-19 group on change in cognitive accuracy between rounds of testing, Round 1 cognitive accuracy was included in models testing association between Round 2 cognitive accuracy and COVID-19 exposures, to control for Round 1 performance and isolate change in accuracy between rounds (Figure 4).

**Figure 4.**
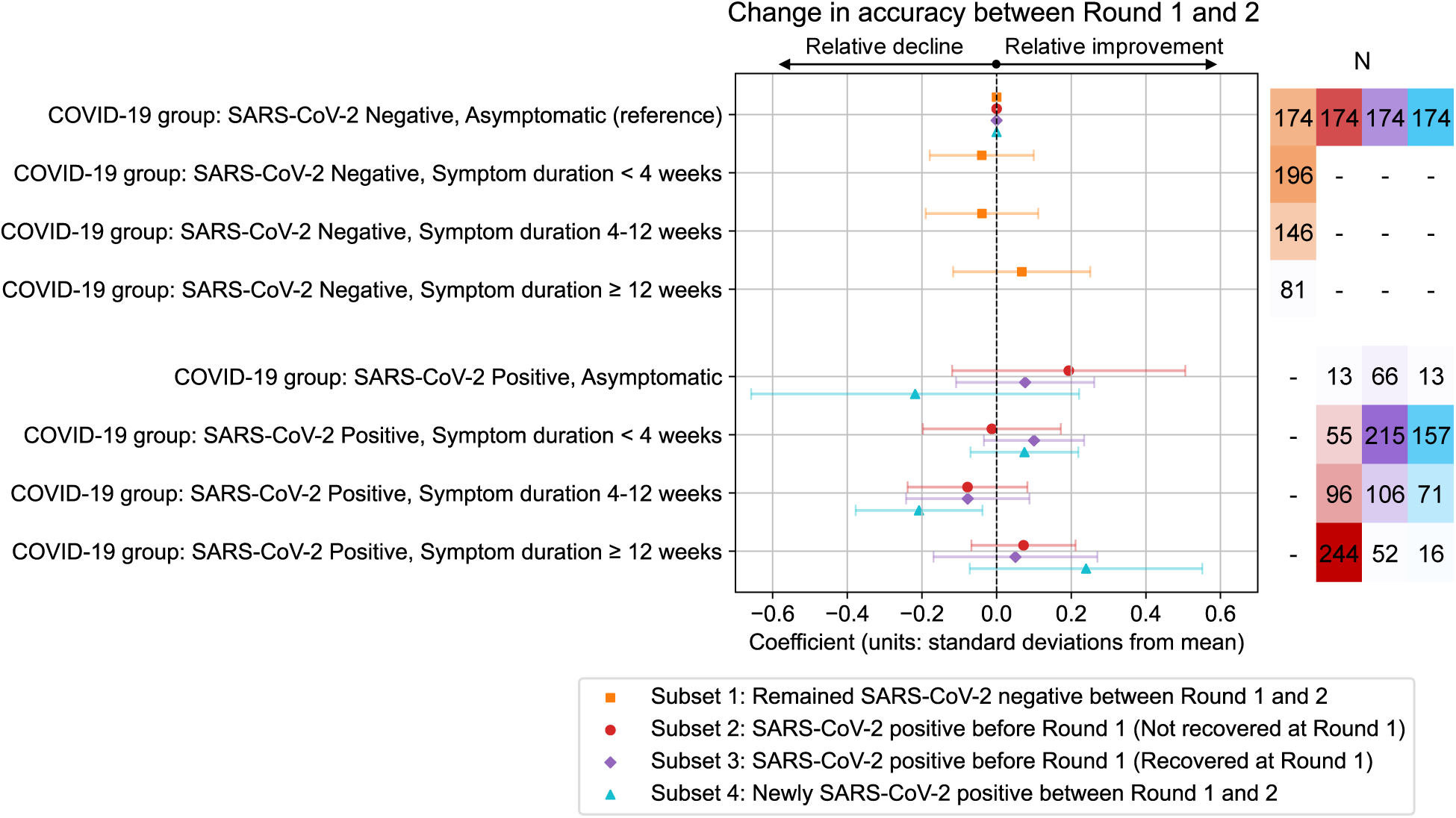
Estimated change in cognitive accuracy between rounds of cognitive testing relative to change observed in reference groups. Standardised coefficients (number of standard deviations from mean) from multivariable ordinary least squares linear regression models testing association between COVID-19 group and cognitive accuracy PCA 1^st^ component standardised scores. Models are presented for subsets of individuals who completed both rounds of testing, selected based on change in COVID-19 group between Round 1 and 2 and self-perceived COVID-19 recovery at Round 1. Models included Round 1 cognitive accuracy score as a mediator in order for coefficients to estimate change in accuracy between rounds due to COVID-19 group, in addition to the following adjustment variables: Age, BMI, Deprivation, Education, Ethnicity, Frailty, Mental health condition count, Physical health condition count, Presentation to hospital, Region, Sex.

Models were run on four subsets of 1768 individuals who completed both rounds of cognitive testing selected based on change in COVID-19 group between Round 1 and 2 and self-perceived COVID-19 recovery at Round 1. Each subset included 174 individuals in the SARS-CoV-2 negative, asymptomatic COVID-19 group at both cognitive assessment rounds to act as the comparison group, in addition to: Subset 1 – 431 individuals who remained SARS-CoV-2 negative between rounds; Subset 2 – 408 individuals who tested SARS-CoV-2 positive *prior* to Round 1 and self-reported as *not recovered* from COVID-19 prior to Round 1; Subset 3 – 439 individuals who tested SARS-CoV-2 positive *prior* to Round 1 and self-reported as *recovered* from COVID-19 prior to Round 1; Subset 4 – 257 individuals who first tested SARS-CoV-2 positive *between* Round 1 and 2. There was no evidence of change in cognitive accuracy between rounds in the Subset 1 individuals who remained SARS-CoV-2 negative (relative to change over time in the negative, asymptomatic reference group). There was also no evidence of change in the individuals with positive tests prior to Round 1 in Subsets 2 and 3, with no differences in change between subsets according to self-perceived recovery from COVID-19. Among Subset 4 individuals who first tested SARS-CoV-2 positive between rounds, only individuals with 4-12 weeks symptom duration showed evidence of change was a relative decline between rounds (β = −0.21 SDs, 95% CI: −0.38, - 0.04, adjusted p = 0.04), with no change evident in the modal group with 0-4 weeks symptom duration.

## Discussion

Our results partially support the hypothesis that those with community-based SARS-CoV-2 infection show cognitive deficits in performance accuracy relative to non-infected individuals, but only among groups with ≥ 12 weeks symptom duration from prospective symptom logging and/or self-reporting as not recovered and “back to normal” following infection. For these individuals with detectable deficits at initial testing, longitudinal follow-up showed deficits persisted at almost two years since infection.

CSSB cohort design with SARS-CoV-2 negative and positive groups across a range of symptom durations enabled effects of infection and symptom duration to be disentangled. At Round 1 testing, lower cognitive task accuracy scores were observed for individuals with positive vs. negative SARS-CoV-2 infection status while controlling for symptom duration, with largest deficits seen for those with ≥ 12 weeks of associated symptoms (Figure 2). Such individuals may self-define as having “long COVID” and meet NICE “Post-COVID-19 syndrome” and WHO “Post COVID-19 condition” definitions [10,11]. The deficits in composite task accuracy scores were comparable in scale to the effect of presentation to hospital during illness, an increase in age of approximately 10 years, or exhibiting mild or moderate symptoms of psychological distress, but smaller than other effects such as lower educational attainment or above threshold fatigue levels (Figure S5). Conversely, we found no evidence of an effect of SARS-CoV-2 infection on average reaction time during tasks (Figure S10) in contrast to observations of individuals who received critical care for COVID-19 using the same assessment platform [7], and a relatively weak effect on variation in reaction time (Figure S9). This is a reassuring finding given the importance of processing speed within cognition and extensive relationships with outcomes such as frailty, dementia and later mortality [43–45].

Importantly, we found no detectable impairment among people who reported as feeling recovered and “back to normal” after their COVID-19 illness, even among individuals who experience long-term symptoms of ≥ 12 weeks (Figure 3). Similarly, presence of ongoing symptoms of psychological distress, fatigue, and functional impairment at the time of cognitive testing partially mediated observed cognitive deficits, suggesting that reductions in these symptoms are elements (but not the whole) of recovery and associated cognitive deficit. These findings are similar to a previous smaller study which found higher cognitive performance for 42 individuals who self-reported as recovered vs. 117 with ongoing symptoms [46]. However, recovery rate was highly correlated with symptom duration (Figure S12), with a recovery rate of only 17% for those with ≥ 12 weeks symptom duration at 38 weeks (IQR: 31-63) since infection. Similarly low recovery rates at 12 months since infection have been reported in other cohorts following individuals with long COVID (15%) and after hospitalisation (29%) [17,47].

Longitudinal follow-up at a follow-up time of 9 months found no evidence of change in cognitive accuracy (neither improvement nor decline) for individuals who had SARS-CoV-2 infection and reported as not recovered before Round 1 (93% prior to vaccination), with deficits persisting at almost two years since infection (median: 84 weeks, IQR: 74, 108).

Conversely, in an opportunistic analysis of individuals who were recruited with SARS-CoV-2 negative statuses but had first infections between rounds of cognitive assessment, we found much less convincing evidence of cognitive sequelae for these later COVID-19 infections. Such infections occurred after vaccination against SARS-CoV-2 (99%) and skewed towards shorter durations than infections before Round 1, which may reflect our sampling strategy, as well as the reduced likelihood of long COVID (illness duration ≥ 4 weeks) for more recent delta vs. alpha and omicron vs. delta variants [48,49], and following vaccination [50].

There are some limitations to our study. Cognitive assessment data prior to SARS-CoV-2 infection was not available for most cases. Despite efforts to address potential selection and participation biases, it is possible some remain. The generalisability of our findings is limited by CSSB cohort composition, which has lower proportions of males, racialised non-white ethnic groups, individuals without university-level education, and those living in more deprived areas than the UK population. As such, replication is needed in other populations. Finally, our study relied on voluntary prospective logging of symptoms and SARS-CoV-2 test results via a smartphone app to derive SARS-CoV-2 infection status and estimate symptom duration as well as retrospective survey responses. Both datasets may be imperfect and incomplete, leading to misclassification in some cases.

In summary, individuals with ≥ 12 weeks symptoms following SARS-CoV-2 infection in the first year of the pandemic had detectable deficits in cognitive accuracy. Those with ongoing symptoms at initial testing did not show cognitive recovery at follow-up 9 months later. The population infected in 2020 with ongoing symptoms, to whom this result is most likely to apply, is sizeable – UK Office for National Statistics estimated that as of January, 2023, 687,000 in the UK were experiencing self-reported long COVID (defined as having ongoing symptoms at more than 4 weeks since infection) after a first infection at least two years previously [13]. The scale of deficits we observed may have detrimental impacts on quality-of-life and daily functioning at an individual level as previously reported [14], as well as socio-economic impacts on society more broadly due to both a reduced capacity to work and an increased need for support. With infrequent and inconsistent identification of long COVID in electronic health care records [51], this work calls for renewed efforts to identify those affected by ongoing symptoms following SARS-CoV-2 infection. Our results highlight the importance of assessing the *ongoing* element of long COVID definitions, which appears to be a better predictor of cognitive impairment due to COVID-19 than symptom duration. Future work needs to focus on trajectories and mechanisms of recovery from ongoing symptoms following COVID-19, as well as the long-term implications on individuals and society of the persistent cognitive deficits observed in this study.

## Supporting information

ICMJE disclosure forms

## Data Availability

Access to data in the CSS Biobank is available to bona fide health researchers on application to the CSS Biobank Management Group. Further details are available online at https://cssbiobank.com/information-for-researchers including application forms and contact information. Analysis code used in this study will be made available openly on Github at https://github.com/nathan-cheetham/CSSBiobank_CognitiveAssessment. Anonymised COVID Symptom Study data are available to researchers to be shared with researchers according to their protocols in the public interest through Health Data Research UK (HDRUK) and Secure Anonymised Information Linkage consortium, housed in the UK Secure Research Platform (Swansea, UK) at https://web.www.healthdatagateway.org/dataset/fddcb382-3051-4394-8436-b92295f14259.

https://github.com/nathan-cheetham/CSSBiobank_CognitiveAssessment

https://web.www.healthdatagateway.org/dataset/fddcb382-3051-4394-8436-b92295f14259

## Contributors

Following CRediT framework https://www.elsevier.com/authors/policies-and-guidelines/credit-author-statement.

Conceptualisation: CJS, RP, NJC, with contribution from all authors

Funding acquisition: CJS, ED, SO

Project administration: VB, PH, AH

Methodology: CJS, RP, NJC, VG, AH, PH, CHS

Formal analysis: NJC

Investigation: NJC, RP, VB, PH, AH, KJD, MHM

Visualisation: NJC

Data curation: VB, LC, LSC, NJC, JD, AH, PH, EK, BM, CHS

Writing – original draft: NJC, CJS, VB, VG, AH, PH, RP, CHS, ED, EM

Writing – review and editing: All authors

## Acknowledgements

We thank COVID Symptom Study Biobank participants, in particular those who completed cognitive testing while experiencing ongoing symptoms. We are grateful to the CSS Biobank Volunteer Advisory Panel for their input on the development of the biobank, and feedback on the design of this study and its significance for patients and the wider public.

We are grateful to staff at ZOE Ltd (including Christina Hu and Joan Capdevila Pujol) for their work on the CSS app, for enabling recruitment to the CSS Biobank and sharing and maintaining CSS data. We are grateful to data and program team staff at TwinsUK (including Andrew Anastasiou, Maria Paz Garcia, Genevieve Lachance, Darioush Yarand) for work on CSSB recruitment, setup and data management. We thank Carl Graham, Jeffrey Seow, Sam Acors, and Neo Kouphou for antibody testing of participant blood samples. We thank Liyuan Chen for work relating to COVID Symptom Study app data processing and management.

The CSS Biobank is supported by the Chronic Disease Research Foundation. Authors affiliated with King’s College London are also supported by the Wellcome Engineering and Physical Sciences Research Council Centre for Medical Engineering at King’s College London (KCL, WT [203148/Z/16/Z]) and the UK Department of Health via the National Institute for Health and Care Research (NIHR) comprehensive Biomedical Research Centre award to Guy’s & St Thomas’ NHS Foundation Trust (GSTT) in partnership with KCL and King’s College Hospital NHS Foundation Trust. Investigators also received support from Medical Research Council, British Heart Foundation, Alzheimer’s Society, European Union, NIHR (including CONVALESCENCE study [COV-LT-0009]), COVID-19 Driver Relief Fund, Wellcome Trust [215010/Z/18/Z], and the NIHR-funded BioResource and Clinical Research Facility. NJC was supported by NIHR [COV-LT-0009]. RSP is a fellow on the Multimorbidity Doctoral Training Programme for Health Professionals, which is supported by the Wellcome Trust [223499/Z/21/Z]. AH and VG are supported by the NIHR Imperial Biomedical Research Centre. WT is supported by the EPSRC Centre for Doctoral Training in Neurotechnology. MHM & KJD are supported by the King’s Together Rapid COVID-19 Call award. MHM is supported by the Wellcome Trust [106223/Z/14/Z]. KJD thanks Fondation Dormeur, Vaduz for funding equipment. SO was supported by the French government, through the 3IA Côte d’Azur Investments in the Future project managed by the National Research Agency (ANR) with the reference number [ANR-19-P3IA-0002]. ZOE Ltd provided in-kind support for all aspects of building, running and supporting the COVID Symptom Study app and service to all users worldwide.

