## Supplemental File 1 for "The effects of COVID-19 on cognitive performance in a community-based cohort: A COVID Symptom Study Biobank observational study"

#### Supplementary information

##### Author list


Nathan J. Cheetham*, 1, https://orcid.org/0000-0002-2259-1556

Rose Penfold, 1,2, https://orcid.org/0000-0001-7023-7108

Valentina Giunchiglia, 3, https://orcid.org/0000-0002-2165-7840

Vicky Bowyer, 1

Carole H. Sudre, 4,5,6, https://orcid.org/0000-0001-5753-428X

Liane S. Canas, 6

Jie Deng, 6

Benjamin Murray, 6

Eric Kerfoot, 6

Michela Antonelli, 6

Khaled Rjoob, 4, https://orcid.org/0000-0002-7598-2323

Erika Molteni, 6, https://orcid.org/0000-0001-7773-8140

Marc F. Österdahl, 1

Nicholas R. Harvey, 1, https://orcid.org/0000-0003-2035-8475

William R. Trender, 3, https://orcid.org/0000-0003-3947-5532

Michael H. Malim, 7

Katie J. Doores, 7

Peter J. Hellyer, 8

Marc Modat, 6

Alexander Hammers, 6,9, https://orcid.org/0000-0001-9530-4848

Sebastien Ourselin, 6, https://orcid.org/0000-0002-5694-5340

Emma L. Duncan, 1,10, https://orcid.org/0000-0002-8143-4403

Adam Hampshire, 3

Claire J. Steves*, 1,10, https://orcid.org/0000-0002-4910-0489

* Corresponding authors

1 Department of Twin Research and Genetic Epidemiology, King's College London, London, United Kingdom

2 Edinburgh Delirium Research Group, Ageing and Health, Usher Institute, University of Edinburgh, Edinburgh, United Kingdom

3 Department of Brain Sciences, Imperial College London, United Kingdom

4 MRC Unit for Lifelong Health and Ageing, Department of Population Health Sciences, University College London, London, United Kingdom

5 Centre for Medical Image Computing, Department of Computer Science, University College London, London, United Kingdom

6 School of Biomedical Engineering & Imaging Sciences, King's College London, London, UK

7 Department of Infectious Diseases, King's College London, London, United Kingdom

8 Centre for Neuroimaging Sciences, King's College London, London, United Kingdom

9 King's College London & Guy's and St Thomas' PET Centre, King's College London, London, United Kingdom

10 Guy's & St Thomas's NHS Foundation Trust, London, United Kingdom

#### S1. Cognitive tasks information

Table S 1. **Description of tasks within cognitive testing.**

| Task order | Task name | Cognition domain(s) tested | Accuracy metric (Transformation method) | Average reaction time metric (Transformation method) | Reaction time variation metric (Transformation method) |
|---|---|---|---|---|---|
| 1 | Immediate memory (words) | Long-term verbal memory encoding and retrieval | Total correct responses (Cube) | Median response time (Log) | Response time interquartile range (Log) |
| 9 | Delayed memory (words) | Long-term verbal memory encoding and retrieval | Total correct responses (Cube) | Median response time (Log) | Response time interquartile range (Log) |
| 2 | Immediate memory (objects) | Long-term visual memory encoding and retrieval | Total correct responses (Cube) | Median response time (Log) | Response time interquartile range (Log) |
| 10 | Delayed memory (objects) | Long-term visual memory encoding and retrieval | Total correct responses (Cube) | Median response time (Log) | Response time interquartile range (Log) |
| 11 | Paired associate learning | Visual working memory | Maximum achieved (Square root) | Median response time (Log) | Response time interquartile range (Log) |
| 12 | Cognitive reflection test | Impulse control, attentional control, processing speed | Proportion of correct responses (Cube) | Median response time (Square root) | Response time interquartile range (Log) |
| 7 | Tower of London | Visual spatial planning | Total correct responses (Square) | Median response time (Square root) | Response time interquartile range (Log) |
| 6 | Spatial span | Visual working memory | Maximum achieved (None) | Median response time (Log) | Response time interquartile range (Log) |
| 5 | Target detection | Visual spatial attention, processing speed | Total correct responses (Cube) | Mean response time (None) | Response time standard deviation (None) |
| 4 | 2D mental manipulations | Spatial mental manipulation | Total correct responses (None) | Median response time (Log) | Response time interquartile range (Log) |
| 3 | Motor control | Motor control, processing speed | Mean distance to target (Log) | Mean response time (Log) | Response time standard deviation (Log) |
| 8 | Verbal analogies | Semantic reasoning | Total correct responses (None) | Median response time (Square root) | Response time interquartile range (Log) |

#### S2. Deriving "COVID-19 group" variable

The following criteria were applied to estimate and validate symptom duration associated with positive and negative tests. To estimate symptom duration, the following criteria must have been met:

- Not a one-time user of app: At least two symptom assessments recorded
- Symptoms reported within time window relevant to test: First "not healthy" assessment is recorded within 14 days either side of antigen test date or earlier than 14 days before antibody test date
- Symptoms reported regularly: Within period of ongoing symptoms, individuals must report as "not healthy" at least once every 14 days
- User reports when symptoms are absent: At least one "healthy" assessment during symptom period
- Absence of symptoms reported after final report of symptoms: To signify likely end of symptoms, at least one "healthy" assessment reported within 14 days following final "not healthy" assessment date [this criterion applied when duration < 28 days only. Once duration exceeds initial acute COVID-19 phase, a more liberal approach was taken to avoid underestimation of symptom duration]

To be considered as "asymptomatic", the following criteria must have been met:

- Not a one-time user of app: At least two symptom assessments recorded
- Absence of symptoms actively reported: One or more 'healthy' assessments in both the 7 days before and after the antigen test date, or earlier than 14 days before antibody test date]
- Presence of symptoms not reported: No 'not healthy' assessments 7 days either side of antigen test date [no 'not healthy' assessments earlier than 14 days before antibody test date]

Where symptom duration estimation criteria were not met, the duration associated with the test was classed as "unknown". Symptom duration was calculated as the number of days between the first date within the test window with reported symptoms and the last date with reported symptoms which met the regularity criteria, plus one day.

For individuals with multiple test results, symptom durations associated with tests were estimated for all positive tests spaced more than 90 days apart, and all negative tests spaced more than 14 days apart. When individuals had multiple test results with varying estimated symptom durations, individuals were assigned to a single "COVID-19 group" by first prioritising positive over negative test results, then taking the test result with the longest estimated symptom duration.

Enzyme-linked immunosorbent assays (ELISA) anti-Spike and anti-Nucleocapsid SARS-CoV-2 antibody tests were also performed internally after recruitment into the CSSB following methods outlined previously [35]. Individuals who reported symptoms but whose first positive SARS-CoV-2 test was a positive ELISA anti-Nucleocapsid (at any time) or anti-Spike (prior to the start of the

UK SARS-CoV-2 vaccination program, 8th December 2020) antibody test were assigned as having "unknown" symptom duration, as it was not possible to associate with confidence the symptom period with the subsequent positive test.

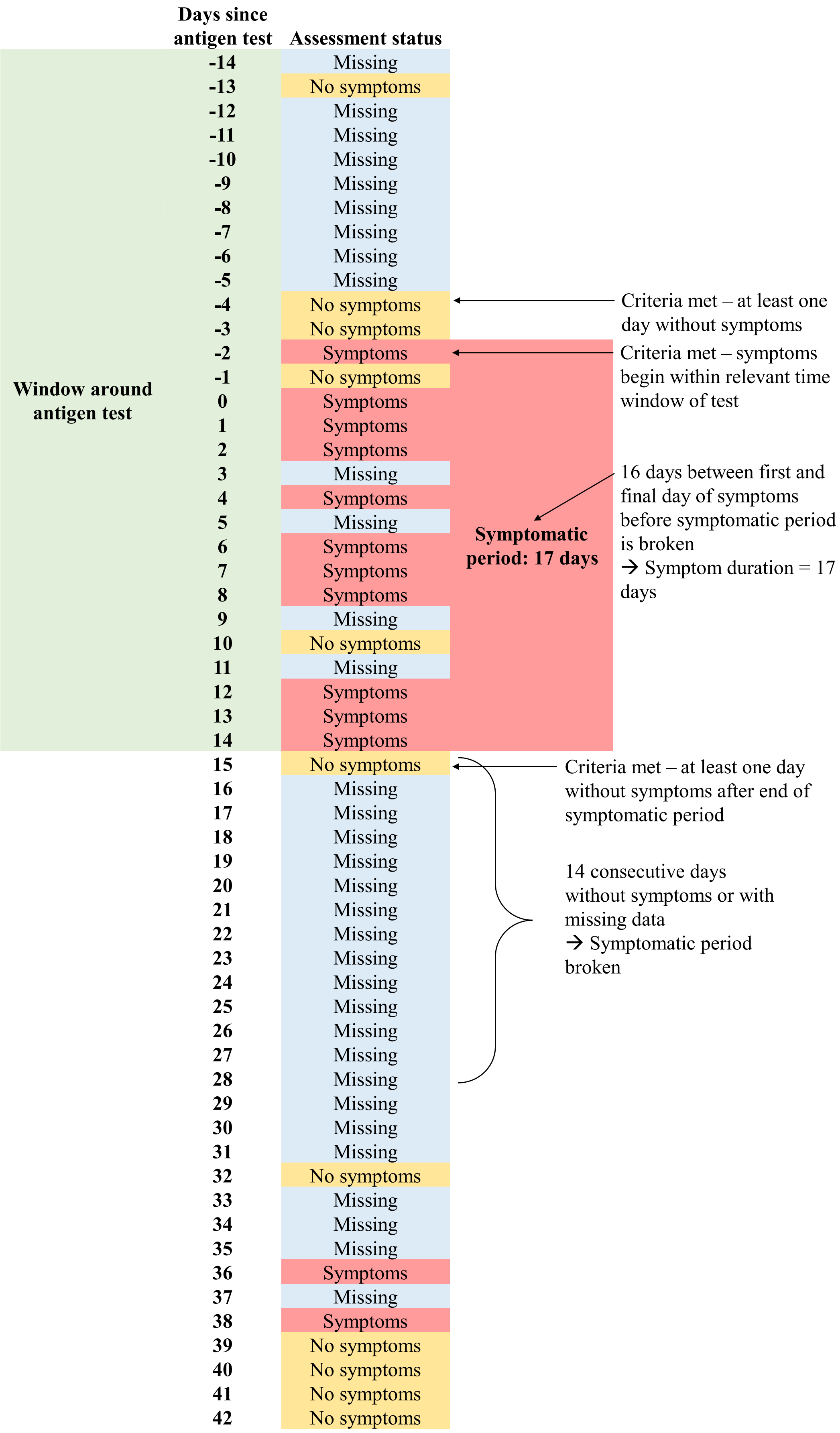


Figure S 1. **Visualisation of symptom duration estimation from symptom reporting.** Hypothetical illustration of how an individual's symptom reporting around a SARS-CoV-2 test is used to estimate symptom duration, highlighting the criteria used in estimations.

#### S3. Generation of inverse participation weights

A forward sequential feature selection approach was taken to find the combination of variables which gave the highest area under the receiver operator curve (AUC-ROC) assessed using a 75:25 train:test data partition. Only variables available prior to cognitive assessment were considered, namely: age, sex, ethnicity, deprivation, region, BMI, physical health condition count, mental health condition count, PHQ-4 category, PRISMA-7 score, COVID-19 group at recruitment, COVID-19 group at invitation to cognitive assessment, presentation to hospital during symptomatic period, and count of prior non-responses to other CSSB studies. Models using all variables had only marginally (< 1%) smaller AUC-ROC values than models which maximised AUC-ROC. Therefore, models using all variables were used to generate weights. From logistic regression models, predicted participation probabilities were generated, inversed and then linearly scaled to sum to the number of individuals invited to cognitive assessment to produce final weights.

#### S4. Multivariable model adjustment variable sets

Table S 2. **Multivariable model adjustment variable sets.** Variables included as adjustments in multivariable models testing associations between outcome and exposures. Adjustment sets were generated from proposed DAGs detailing participation in cognitive assessment and cognitive performance. Highest educational attainment was not included as an adjustment variable in cognitive performance models of all Round 1 participants, due to missing data.

| | Adjustment set | |
|---|---|---|
| Exposure | Outcome: Participation in cognitive assessment | Outcome: Cognitive performance |
| Age | Ethnicity, Sex | Ethnicity, Sex |
| Sex | Age, Ethnicity | Age, Ethnicity |
| Ethnicity | Age, Sex | Age, Sex |
| Highest educational attainment | - | Age |
| Region | Age, BMI, Deprivation, Ethnicity, Frailty (PRISMA-7), Mental health condition count, Physical health condition count, Sex | Age, BMI, Deprivation, Education, Ethnicity, Frailty (PRISMA-7), Mental health condition count, Physical health condition count, Sex |
| Local area deprivation (IMD) | Age, BMI, Ethnicity, Frailty (PRISMA-7), Mental health condition count, Physical health condition count, Region, Sex | Age, BMI, Education, Ethnicity, Frailty (PRISMA-7), Mental health condition count, Physical health condition count, Region, Sex |
| Physical health condition count | Age, BMI, Deprivation, Ethnicity, Frailty (PRISMA-7), Mental health condition count, Region, Sex | Age, BMI, Deprivation, Education, Ethnicity, Frailty (PRISMA-7), Mental health condition count, Region, Sex |
| Mental health condition count | Age, BMI, Deprivation, Ethnicity, Frailty (PRISMA-7), Physical health condition count, Region, Sex | Age, BMI, Deprivation, Education, Ethnicity, Frailty (PRISMA-7), Physical health condition count, Region, Sex |
| BMI | Age, Deprivation, Ethnicity, Frailty (PRISMA-7), Mental health condition count, Physical health condition count, Region, Sex | Age, Deprivation, Education, Ethnicity, Frailty (PRISMA-7), Mental health condition count, Physical health condition count, Region, Sex |

| | | |
|---|---|---|
| Frailty (PRISMA-7) | Age, BMI, Deprivation, Ethnicity, Mental health condition count, Physical health condition count, Region, Sex | Age, BMI, Deprivation, Education, Ethnicity, Mental health condition count, Physical health condition count, Region, Sex |
| COVID-19 group at recruitment | Age, BMI, Deprivation, Ethnicity, Frailty (PRISMA-7), Mental health condition count, Physical health condition count, Region, Sex | - |
| COVID-19 group at invitation | Age, COVID-19 group at recruitment, Deprivation, Ethnicity, Mental health condition count, Mental health in prior CSSB studies (PHQ-4), Number of non-responses in prior CSSB studies, Presentation to hospital, Region, Sex | Age, BMI, Deprivation, Education, Ethnicity, Frailty (PRISMA-7), Mental health condition count, Physical health condition count, Presentation to hospital, Region, Sex |
| SARS-CoV-2 test result at invitation | - | Age, BMI, Deprivation, Education, Ethnicity, Frailty (PRISMA-7), Mental health condition count, Physical health condition count, Presentation to hospital, Region, Sex, Symptom duration |
| Presentation to hospital | Age, COVID-19 group at invitation, COVID-19 group at recruitment, Deprivation, Ethnicity, Mental health condition count, Mental health in prior CSSB studies (PHQ-4), Number of non-responses in prior CSSB studies, Region, Sex | Age, BMI, Deprivation, Education, Ethnicity, Frailty (PRISMA-7), Mental health condition count, Physical health condition count, Presentation to hospital, Region, Sex, Symptom duration |
| Mental health in prior CSSB studies (PHQ-4) | Age, COVID-19 group at invitation, COVID-19 group at recruitment, Deprivation, Ethnicity, Mental health condition count, Number of non-responses in prior CSSB studies, Presentation to hospital, Region, Sex | Age, BMI, COVID-19 group at invitation, Deprivation, Education, Ethnicity, Frailty (PRISMA-7), Mental health condition count, Physical health condition count, Region, Sex |
| Number of non-responses in prior CSSB studies | Age, COVID-19 group at invitation, COVID-19 group at recruitment, Deprivation, Ethnicity, Mental health condition count, Mental health in prior CSSB studies (PHQ-4), Presentation to hospital, Region, Sex | - |
| Ongoing symptoms (WSAS/CFS/PHQ-4) | - | Age, BMI, COVID-19 group at invitation, Education, Ethnicity, Frailty (PRISMA-7), Mental health condition count, Physical health condition count, Presentation to hospital, Sex |

### S5. Directed acyclic graph model code

#### Participation outcome graph:

*dag {*

*bb="-2.632,-3.208,2.645,3.17"*

*"COVID-19 group at invitation" [pos="0.696,-1.391"]*

*"COVID-19 group at recruitment" [pos="-0.348,-0.881"]*

*"Frailty (PRISMA-7)" [pos="-1.324,1.280"]*

*"Mental health condition count" [pos="-1.220,-0.364"]*

*"Mental health in prior CSSB studies (PHQ-4)" [pos="0.517,0.750"]*

*"Number of non-responses in prior CSSB studies" [pos="0.700,-2.320"]*

*"Physical health condition count" [pos="-1.115,0.140"]*

*"Presentation to hospital" [pos="0.681,-0.331"]*

*Age [pos="-2.095,-1.867"]*

*BMI [pos="-1.123,0.564"]*

*Deprivation [pos="-1.100,-2.048"]*

*Ethnicity [pos="-2.103,-0.682"]*

*Participation [outcome,pos="1.721,-0.907"]*

*Region [pos="-1.216,-2.760"]*

*Sex [pos="-2.233,-1.299"]*

*"COVID-19 group at invitation" -> Participation*

*"COVID-19 group at invitation" <-> "Mental health in prior CSSB studies (PHQ-4)"*

*"COVID-19 group at invitation" <-> "Number of non-responses in prior CSSB studies"*

*"COVID-19 group at invitation" <-> "Presentation to hospital"*

*"COVID-19 group at recruitment" -> "COVID-19 group at invitation"*

*"COVID-19 group at recruitment" -> "Mental health in prior CSSB studies (PHQ-4)"*

*"COVID-19 group at recruitment" -> "Number of non-responses in prior CSSB studies"*

*"COVID-19 group at recruitment" -> "Presentation to hospital"*

*"COVID-19 group at recruitment" -> Participation*

*"Frailty (PRISMA-7)" -> "COVID-19 group at invitation"*

*"Frailty (PRISMA-7)" -> "COVID-19 group at recruitment"*

*"Frailty (PRISMA-7)" -> "Mental health in prior CSSB studies (PHQ-4)"*

*"Frailty (PRISMA-7)" -> "Presentation to hospital"*

*"Frailty (PRISMA-7)" <-> "Mental health condition count"*

*"Frailty (PRISMA-7)" <-> "Physical health condition count"*

*"Frailty (PRISMA-7)" <-> BMI*

*"Frailty (PRISMA-7)" <-> Deprivation*

*"Frailty (PRISMA-7)" <-> Region*

*"Mental health condition count" -> "COVID-19 group at invitation"*

*"Mental health condition count" -> "COVID-19 group at recruitment"*

*"Mental health condition count" -> "Mental health in prior CSSB studies (PHQ-4)"*

*"Mental health condition count" -> "Presentation to hospital"*

*"Mental health condition count" -> Participation*

*"Mental health condition count" <-> "Physical health condition count"*

*"Mental health condition count" <-> BMI*

*"Mental health condition count" <-> Deprivation*

*"Mental health condition count" <-> Region*

*"Mental health in prior CSSB studies (PHQ-4)" -> Participation*

*"Mental health in prior CSSB studies (PHQ-4)" <-> "Presentation to hospital"*

*"Number of non-responses in prior CSSB studies" -> Participation*

*"Physical health condition count" -> "COVID-19 group at invitation"*

*"Physical health condition count" -> "COVID-19 group at recruitment"*

*"Physical health condition count" -> "Mental health in prior CSSB studies (PHQ-4)"*

*"Physical health condition count" -> "Presentation to hospital"*

*"Physical health condition count" <-> BMI*

*"Physical health condition count" <-> Deprivation*

*"Presentation to hospital" -> Participation*

*Age -> "COVID-19 group at invitation"*

*Age -> "COVID-19 group at recruitment"*

*Age -> "Frailty (PRISMA-7)"*

*Age -> "Mental health condition count"*

*Age -> "Mental health in prior CSSB studies (PHQ-4)"*

*Age -> "Number of non-responses in prior CSSB studies"*

*Age -> "Physical health condition count"*

*Age -> "Presentation to hospital"*

*Age -> BMI*

*Age -> Deprivation*

*Age -> Participation*

*Age -> Region*

*Age <-> Ethnicity*

*Age <-> Sex*

*BMI -> "COVID-19 group at invitation"*

*BMI -> "COVID-19 group at recruitment"*

*BMI -> "Mental health in prior CSSB studies (PHQ-4)"*

*BMI -> "Number of non-responses in prior CSSB studies"*

*BMI -> "Presentation to hospital"*

*BMI <-> Deprivation*

*BMI <-> Region*

*Deprivation -> "COVID-19 group at invitation"*

*Deprivation -> "COVID-19 group at recruitment"*

*Deprivation -> "Mental health in prior CSSB studies (PHQ-4)"*

*Deprivation -> "Number of non-responses in prior CSSB studies"*

*Deprivation -> "Presentation to hospital"*

*Deprivation -> Participation*

*Deprivation <-> Region*

*Ethnicity -> "COVID-19 group at invitation"*

*Ethnicity -> "COVID-19 group at recruitment"*

*Ethnicity -> "Mental health condition count"*

*Ethnicity -> "Mental health in prior CSSB studies (PHQ-4)"*

*Ethnicity -> "Number of non-responses in prior CSSB studies"*

*Ethnicity -> "Physical health condition count"*

*Ethnicity -> "Presentation to hospital"*

*Ethnicity -> BMI*

*Ethnicity -> Deprivation*

*Ethnicity -> Participation*

*Ethnicity -> Region*

*Region -> "COVID-19 group at invitation"*

*Region -> "COVID-19 group at recruitment"*

*Region -> "Mental health in prior CSSB studies (PHQ-4)"*

*Region -> "Number of non-responses in prior CSSB studies"*

*Region -> Participation*

*Sex -> "COVID-19 group at invitation"*

*Sex -> "COVID-19 group at recruitment"*

*Sex -> "Frailty (PRISMA-7)"*

*Sex -> "Mental health condition count"*

*Sex -> "Mental health in prior CSSB studies (PHQ-4)"*

*Sex -> "Physical health condition count"*

*Sex -> BMI*

*Sex -> Deprivation*

*Sex -> Participation*

*}*

#### Cognitive performance outcome graph:

*dag {*

*bb="-2.632,-3.208,2.645,3.17"*

*"COVID-19 group at invitation" [pos="0.081,-0.531"]*

*"Cognitive performance" [outcome,pos="1.948,0.034"]*

*"Frailty (PRISMA-7)" [pos="-0.869,1.877"]*

*"Mental health condition count" [pos="-0.798,0.126"]*

*"Ongoing symptoms (WSAS/CFS/PHQ-4)" [pos="0.983,-1.531"]*

*"Physical health condition count" [pos="-0.858,0.651"]*

*"Presentation to hospital" [pos="0.293,0.707"]*

*Age [pos="-2.095,-1.867"]*

*BMI [pos="-0.972,1.205"]*

*Deprivation [pos="-0.773,-2.205"]*

*Education [pos="-1.428,-2.121"]*

*Ethnicity [pos="-2.092,-0.711"]*

*Region [pos="-0.870,-2.717"]*

*Sex [pos="-2.222,-1.292"]*

*"COVID-19 group at invitation" -> "Cognitive performance"*

*"COVID-19 group at invitation" -> "Ongoing symptoms (WSAS/CFS/PHQ-4)"*

*"COVID-19 group at invitation" <-> "Presentation to hospital"*

*"Frailty (PRISMA-7)" -> "COVID-19 group at invitation"*

*"Frailty (PRISMA-7)" -> "Cognitive performance"*

*"Frailty (PRISMA-7)" -> "Ongoing symptoms (WSAS/CFS/PHQ-4)"*

*"Frailty (PRISMA-7)" -> "Presentation to hospital"*

*"Frailty (PRISMA-7)" <-> "Mental health condition count"*

*"Frailty (PRISMA-7)" <-> "Physical health condition count"*

*"Frailty (PRISMA-7)" <-> BMI*

*"Frailty (PRISMA-7)" <-> Deprivation*

*"Frailty (PRISMA-7)" <-> Region*

*"Mental health condition count" -> "COVID-19 group at invitation"*

*"Mental health condition count" -> "Cognitive performance"*

*"Mental health condition count" -> "Ongoing symptoms (WSAS/CFS/PHQ-4)"*

*"Mental health condition count" -> "Presentation to hospital"*

*"Mental health condition count" <-> "Physical health condition count"*

*"Mental health condition count" <-> BMI*

*"Mental health condition count" <-> Deprivation*

*"Mental health condition count" <-> Region*

*"Ongoing symptoms (WSAS/CFS/PHQ-4)" -> "Cognitive performance"*

*"Physical health condition count" -> "COVID-19 group at invitation"*

*"Physical health condition count" -> "Cognitive performance"*

*"Physical health condition count" -> "Ongoing symptoms (WSAS/CFS/PHQ-4)"*

*"Physical health condition count" -> "Presentation to hospital"*

*"Physical health condition count" <-> BMI*

*"Physical health condition count" <-> Deprivation*

*"Presentation to hospital" -> "Cognitive performance"*

*"Presentation to hospital" -> "Ongoing symptoms (WSAS/CFS/PHQ-4)"*

*Age -> "COVID-19 group at invitation"*

*Age -> "Cognitive performance"*

*Age -> "Frailty (PRISMA-7)"*

*Age -> "Mental health condition count"*

*Age -> "Ongoing symptoms (WSAS/CFS/PHQ-4)"*

*Age -> "Physical health condition count"*

*Age -> "Presentation to hospital"*

*Age -> BMI*

*Age -> Deprivation*

*Age -> Education*

*Age -> Region*

*Age <-> Ethnicity*

*Age <-> Sex*

*BMI -> "COVID-19 group at invitation"*

*BMI -> "Cognitive performance"*

*BMI -> "Ongoing symptoms (WSAS/CFS/PHQ-4)"*

*BMI -> "Presentation to hospital"*

*BMI <-> Deprivation*

*BMI <-> Region*

*Deprivation -> "COVID-19 group at invitation"*

*Deprivation -> "Ongoing symptoms (WSAS/CFS/PHQ-4)"*

*Deprivation <-> Region*

*Education -> "COVID-19 group at invitation"*

*Education -> "Cognitive performance"*

*Education -> "Ongoing symptoms (WSAS/CFS/PHQ-4)"*

*Education -> Deprivation*

*Education -> Region*

*Ethnicity -> "COVID-19 group at invitation"*

*Ethnicity -> "Cognitive performance"*

*Ethnicity -> "Ongoing symptoms (WSAS/CFS/PHQ-4)"*

*Ethnicity -> "Presentation to hospital"*

*Ethnicity -> Deprivation*

*Ethnicity -> Region*

*Region -> "COVID-19 group at invitation"*

*Region -> "Ongoing symptoms (WSAS/CFS/PHQ-4)"*

*Sex -> "COVID-19 group at invitation"*

*Sex -> "Cognitive performance"*

*Sex -> "Frailty (PRISMA-7)"*

*Sex -> "Mental health condition count"*

*Sex -> "Ongoing symptoms (WSAS/CFS/PHQ-4)"*

*Sex -> "Physical health condition count"*

*Sex -> BMI*

*Sex -> Deprivation*

*}*

### S6. Sample selection

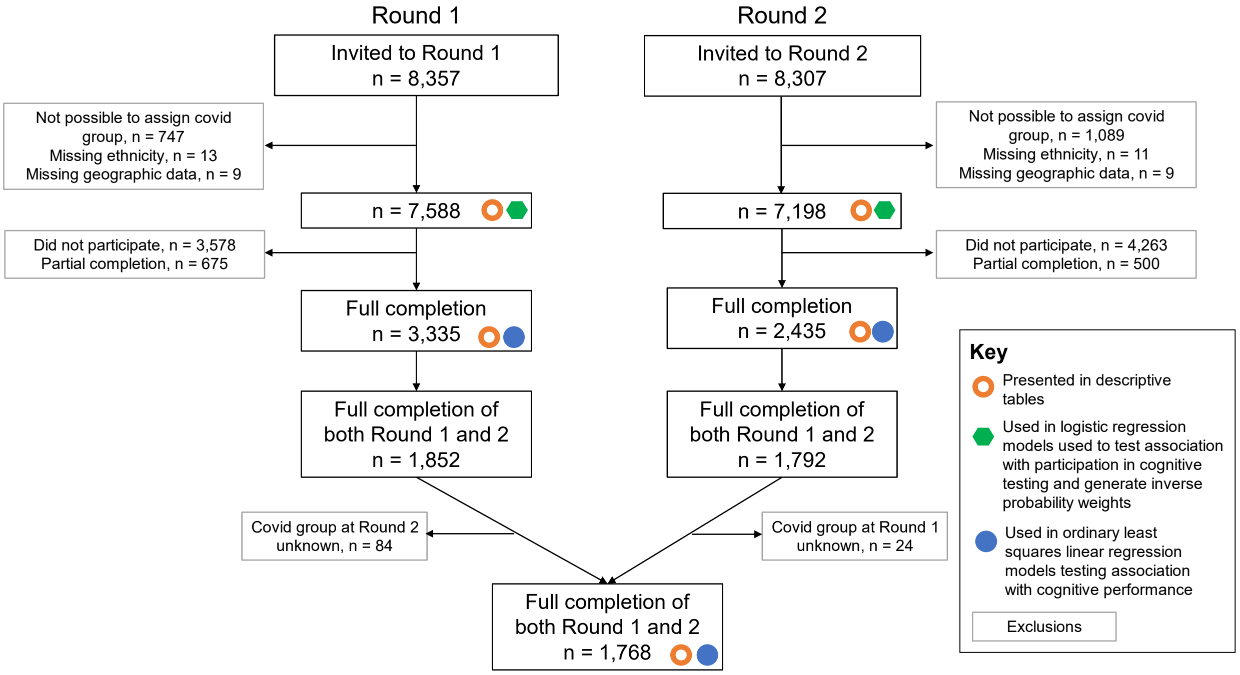


Figure S 2. **Cognitive testing participation flowchart.** Full and partial completion refer to completion of all 12 individual cognitive tasks or between 1 and 11 tasks respectively.

#### S7. Additional sample characteristics with split by COVID-19 group

Table S 3. **Round 1 sample characteristics, split by COVID-19 group**. Additional sample characteristics for COVID Symptom Study Biobank participants who participated in Round 1 of cognitive assessment, with additional splits by COVID-19 group. Counts and proportions are presented except for weeks between cognitive assessment and symptom start/test date, where median and interquartile range is presented. Highest educational attainment level, PHQ-4, Chalder Fatigue Scale and WSAS were collected as part of Round 2 of cognitive assessment. Other variables were collected or derived from data collected at recruitment or prior to analyses.

| | | | COVID-19 group: SARS-CoV-2 result, symptom duration | | | | | | | |
|---|---|---|---|---|---|---|---|---|---|---|
| Variable | Category | Round 1 (full completion) | Negative, asymptomatic | Negative, < 4 weeks | Negative, 4-12 weeks | Negative, >= 12 weeks | Positive, asymptomatic | Positive, < 4 weeks | Positive, 4-12 weeks | Positive, >= 12 weeks |
| N | | 3335 | 641 | 379 | 372 | 206 | 256 | 589 | 397 | 495 |
| Age group (years) | 18-30 | 43 (1.3%) | 7 (1.1%) | 8 (2.1%) | < 5 | < 5 | < 5 | 9 (1.5%) | 7 (1.8%) | < 5 |
| Age group (years) | 30-40 | 214 (6.4%) | 40 (6.2%) | 29 (7.7%) | 28 (7.5%) | 7 (3.4%) | 11 (4.3%) | 39 (6.6%) | 33 (8.3%) | 27 (5.5%) |
| Age group (years) | 40-50 | 539 (16.2%) | 90 (14.0%) | 67 (17.7%) | 67 (18.0%) | 33 (16.0%) | 28 (10.9%) | 80 (13.6%) | 78 (19.6%) | 96 (19.4%) |
| Age group (years) | 50-60 (reference) | 1183 (35.5%) | 204 (31.8%) | 150 (39.6%) | 138 (37.1%) | 79 (38.3%) | 81 (31.6%) | 200 (34.0%) | 144 (36.3%) | 187 (37.8%) |
| Age group (years) | 60-70 | 1042 (31.2%) | 231 (36.0%) | 102 (26.9%) | 109 (29.3%) | 65 (31.6%) | 95 (37.1%) | 194 (32.9%) | 104 (26.2%) | 142 (28.7%) |
| Age group (years) | 70-80 | 293 (8.8%) | 63 (9.8%) | 22 (5.8%) | 24 (6.5%) | 20 (9.7%) | 38 (14.8%) | 64 (10.9%) | 29 (7.3%) | 33 (6.7%) |
| Age group (years) | ≥ 80 | 21 (0.6%) | 6 (0.9%) | < 5 | < 5 | < 5 | 0 | < 5 | < 5 | 6 (1.2%) |
| Sex | Female (reference) | 2697 (80.9%) | 523 (81.6%) | 330 (87.1%) | 306 (82.3%) | 157 (76.2%) | 207 (80.9%) | 453 (76.9%) | 320 (80.6%) | 401 (81.0%) |
| Sex | Male | 638 (19.1%) | 118 (18.4%) | 49 (12.9%) | 66 (17.7%) | 49 (23.8%) | 49 (19.1%) | 136 (23.1%) | 77 (19.4%) | 94 (19.0%) |
| Ethnicity | Asian/Asian British | 25 (0.7%) | < 5 | < 5 | 5 (1.3%) | < 5 | < 5 | < 5 | 9 (2.3%) | 0 |
| Ethnicity | Black/Black British | 12 (0.4%) | < 5 | 0 | < 5 | 0 | < 5 | < 5 | < 5 | < 5 |
| Ethnicity | Mixed/Multiple | 41 (1.2%) | 7 (1.1%) | < 5 | 10 (2.7%) | < 5 | 0 | < 5 | 8 (2.0%) | 7 (1.4%) |
| Ethnicity | Other | 44 (1.3%) | < 5 | < 5 | 6 (1.6%) | < 5 | 10 (3.9%) | 7 (1.2%) | < 5 | 10 (2.0%) |
| Ethnicity | White (reference) | 3213 (96.3%) | 628 (98.0%) | 368 (97.1%) | 349 (93.8%) | 200 (97.1%) | 244 (95.3%) | 573 (97.3%) | 375 (94.5%) | 476 (96.2%) |
| Highest educational attainment level | Data not available | 1194 (35.8%) | 200 (31.2%) | 145 (38.3%) | 120 (32.3%) | 63 (30.6%) | 109 (42.6%) | 255 (43.3%) | 155 (39.0%) | 147 (29.7%) |
| Highest educational attainment level | Postgraduate degree or higher | 658 (30.7%) | 121 (27.4%) | 77 (32.9%) | 91 (36.1%) | 55 (38.5%) | 40 (27.2%) | 103 (30.8%) | 72 (29.8%) | 99 (28.4%) |

| | | | | | | | | | | |
|---|---|---|---|---|---|---|---|---|---|---|
| Highest educational attainment level | Undergraduate degree (reference) | 794 (37.1%) | 162 (36.7%) | 89 (38.0%) | 87 (34.5%) | 52 (36.4%) | 58 (39.5%) | 114 (34.1%) | 90 (37.2%) | 142 (40.8%) |
| Highest educational attainment level | Less than undergraduate degree | 661 (30.9%) | 153 (34.7%) | 65 (27.8%) | 70 (27.8%) | 36 (25.2%) | 47 (32.0%) | 111 (33.2%) | 75 (31.0%) | 104 (29.9%) |
| Highest educational attainment level | Other/Prefer not to say | 28 (1.3%) | 5 (1.1%) | < 5 | < 5 | 0 | < 5 | 6 (1.8%) | 5 (2.1%) | < 5 |
| Region | East Midlands | 192 (5.8%) | 35 (5.5%) | 17 (4.5%) | 21 (5.6%) | 12 (5.8%) | 14 (5.5%) | 31 (5.3%) | 23 (5.8%) | 39 (7.9%) |
| Region | East of England | 382 (11.5%) | 70 (10.9%) | 48 (12.7%) | 48 (12.9%) | 23 (11.2%) | 32 (12.5%) | 60 (10.2%) | 53 (13.4%) | 48 (9.7%) |
| Region | London (reference) | 617 (18.5%) | 93 (14.5%) | 65 (17.2%) | 56 (15.1%) | 31 (15.0%) | 65 (25.4%) | 137 (23.3%) | 81 (20.4%) | 89 (18.0%) |
| Region | North East | 103 (3.1%) | 30 (4.7%) | 6 (1.6%) | 8 (2.2%) | 7 (3.4%) | 7 (2.7%) | 17 (2.9%) | 14 (3.5%) | 14 (2.8%) |
| Region | North West | 305 (9.1%) | 48 (7.5%) | 25 (6.6%) | 24 (6.5%) | 11 (5.3%) | 25 (9.8%) | 74 (12.6%) | 43 (10.8%) | 55 (11.1%) |
| Region | Northern Ireland | 11 (0.3%) | < 5 | < 5 | < 5 | 0 | 0 | < 5 | < 5 | < 5 |
| Region | Scotland | 123 (3.7%) | 31 (4.8%) | 25 (6.6%) | 17 (4.6%) | 11 (5.3%) | < 5 | 9 (1.5%) | 13 (3.3%) | 14 (2.8%) |
| Region | South East | 657 (19.7%) | 143 (22.3%) | 81 (21.4%) | 77 (20.7%) | 49 (23.8%) | 50 (19.5%) | 115 (19.5%) | 66 (16.6%) | 76 (15.4%) |
| Region | South West | 354 (10.6%) | 67 (10.5%) | 48 (12.7%) | 53 (14.2%) | 26 (12.6%) | 20 (7.8%) | 48 (8.1%) | 39 (9.8%) | 53 (10.7%) |
| Region | Wales | 161 (4.8%) | 36 (5.6%) | 17 (4.5%) | 18 (4.8%) | 17 (8.3%) | 11 (4.3%) | 16 (2.7%) | 17 (4.3%) | 29 (5.9%) |
| Region | West Midlands | 221 (6.6%) | 48 (7.5%) | 30 (7.9%) | 23 (6.2%) | 6 (2.9%) | 17 (6.6%) | 40 (6.8%) | 25 (6.3%) | 32 (6.5%) |
| Region | Yorkshire and The Humber | 209 (6.3%) | 38 (5.9%) | 15 (4.0%) | 26 (7.0%) | 13 (6.3%) | 12 (4.7%) | 41 (7.0%) | 22 (5.5%) | 42 (8.5%) |
| Local area deprivation (IMD) | Quintile 1 (most 20% deprived areas) | 188 (5.6%) | 37 (5.8%) | 22 (5.8%) | 17 (4.6%) | 12 (5.8%) | 11 (4.3%) | 33 (5.6%) | 25 (6.3%) | 31 (6.3%) |
| Local area deprivation (IMD) | Quintile 2 | 413 (12.4%) | 82 (12.8%) | 35 (9.2%) | 53 (14.2%) | 22 (10.7%) | 19 (7.4%) | 69 (11.7%) | 50 (12.6%) | 83 (16.8%) |
| Local area deprivation (IMD) | Quintile 3 (reference) | 682 (20.4%) | 116 (18.1%) | 81 (21.4%) | 81 (21.8%) | 43 (20.9%) | 51 (19.9%) | 120 (20.4%) | 83 (20.9%) | 107 (21.6%) |
| Local area deprivation (IMD) | Quintile 4 | 883 (26.5%) | 162 (25.3%) | 117 (30.9%) | 98 (26.3%) | 52 (25.2%) | 69 (27.0%) | 157 (26.7%) | 111 (28.0%) | 117 (23.6%) |
| Local area deprivation (IMD) | Quintile 5 (least 20% deprived areas) | 1169 (35.1%) | 244 (38.1%) | 124 (32.7%) | 123 (33.1%) | 77 (37.4%) | 106 (41.4%) | 210 (35.7%) | 128 (32.2%) | 157 (31.7%) |
| BMI | Data not available | 8 (0.2%) | < 5 | < 5 | < 5 | < 5 | < 5 | < 5 | < 5 | < 5 |
| BMI | < 18.5 kg/m^2 | 33 (1.0%) | 10 (1.6%) | < 5 | < 5 | < 5 | < 5 | < 5 | < 5 | 8 (1.6%) |
| BMI | 18.5-25 kg/m^2 (reference) | 1434 (43.1%) | 296 (46.2%) | 170 (45.0%) | 157 (42.4%) | 76 (36.9%) | 118 (46.3%) | 260 (44.3%) | 171 (43.2%) | 186 (37.6%) |
| BMI | 25-30 kg/m^2 | 1056 (31.7%) | 196 (30.6%) | 112 (29.6%) | 121 (32.7%) | 63 (30.6%) | 84 (32.9%) | 202 (34.4%) | 131 (33.1%) | 147 (29.7%) |

| | | | | | | | | | | |
|---|---|---|---|---|---|---|---|---|---|---|
| BMI | ≥ 30 kg/m^2 | 804 (24.2%) | 138 (21.6%) | 95 (25.1%) | 90 (24.3%) | 63 (30.6%) | 51 (20.0%) | 122 (20.8%) | 91 (23.0%) | 154 (31.1%) |
| Physical health conditions | None (reference) | 2564 (76.9%) | 542 (84.6%) | 292 (77.0%) | 261 (70.2%) | 136 (66.0%) | 214 (83.6%) | 448 (76.1%) | 296 (74.6%) | 375 (75.8%) |
| Physical health conditions | One | 673 (20.2%) | 92 (14.4%) | 80 (21.1%) | 92 (24.7%) | 56 (27.2%) | 38 (14.8%) | 122 (20.7%) | 91 (22.9%) | 102 (20.6%) |
| Physical health conditions | Two or more | 98 (2.9%) | 7 (1.1%) | 7 (1.8%) | 19 (5.1%) | 14 (6.8%) | < 5 | 19 (3.2%) | 10 (2.5%) | 18 (3.6%) |
| Mental health conditions | Data not available | 533 (16.0%) | 59 (9.2%) | 55 (14.5%) | 59 (15.9%) | 20 (9.7%) | 52 (20.3%) | 118 (20.0%) | 94 (23.7%) | 76 (15.4%) |
| Mental health conditions | None (reference) | 2076 (74.1%) | 473 (81.3%) | 242 (74.7%) | 196 (62.6%) | 120 (64.5%) | 175 (85.8%) | 375 (79.6%) | 208 (68.6%) | 287 (68.5%) |
| Mental health conditions | One | 502 (17.9%) | 84 (14.4%) | 54 (16.7%) | 86 (27.5%) | 31 (16.7%) | 27 (13.2%) | 72 (15.3%) | 60 (19.8%) | 88 (21.0%) |
| Mental health conditions | Two | 163 (5.8%) | 20 (3.4%) | 22 (6.8%) | 22 (7.0%) | 20 (10.8%) | < 5 | 19 (4.0%) | 23 (7.6%) | 35 (8.4%) |
| Mental health conditions | Three or more | 61 (2.2%) | 5 (0.9%) | 6 (1.9%) | 9 (2.9%) | 15 (8.1%) | 0 | 5 (1.1%) | 12 (4.0%) | 9 (2.1%) |
| PRISMA-7 | Score 0/7 | 700 (21.0%) | 138 (21.5%) | 85 (22.4%) | 63 (16.9%) | 38 (18.4%) | 70 (27.3%) | 125 (21.2%) | 73 (18.4%) | 108 (21.8%) |
| PRISMA-7 | Score 1/7 | 1855 (55.6%) | 369 (57.6%) | 232 (61.2%) | 214 (57.5%) | 89 (43.2%) | 137 (53.5%) | 328 (55.7%) | 226 (56.9%) | 260 (52.5%) |
| PRISMA-7 | Score 2/7 | 613 (18.4%) | 112 (17.5%) | 45 (11.9%) | 66 (17.7%) | 47 (22.8%) | 45 (17.6%) | 113 (19.2%) | 82 (20.7%) | 103 (20.8%) |
| PRISMA-7 | Score 3/7 | 104 (3.1%) | 17 (2.7%) | 11 (2.9%) | 20 (5.4%) | 16 (7.8%) | < 5 | 17 (2.9%) | 9 (2.3%) | 12 (2.4%) |
| PRISMA-7 | Score 4/7 | 38 (1.1%) | < 5 | < 5 | 7 (1.9%) | 10 (4.9%) | < 5 | 5 (0.8%) | 5 (1.3%) | < 5 |
| PRISMA-7 | Score 5/7 | 21 (0.6%) | < 5 | < 5 | 0 | 5 (2.4%) | 0 | < 5 | < 5 | 8 (1.6%) |
| PRISMA-7 | Score 6/7 | < 5 | < 5 | 0 | < 5 | < 5 | 0 | 0 | 0 | 0 |
| Presented to hospital during symptomatic period | No (reference) | 3054 (91.6%) | 637 (99.4%) | 369 (97.4%) | 350 (94.1%) | 174 (84.5%) | 253 (98.8%) | 569 (96.6%) | 339 (85.4%) | 363 (73.3%) |
| Presented to hospital during symptomatic period | Yes | 281 (8.4%) | < 5 | 10 (2.6%) | 22 (5.9%) | 32 (15.5%) | < 5 | 20 (3.4%) | 58 (14.6%) | 132 (26.7%) |
| Self-perceived COVID-19 recovery | N/A, no self-reported COVID-19 (reference) | 1279 (38.4%) | 578 (90.2%) | 319 (84.2%) | 267 (71.8%) | 115 (55.8%) | 0 | 0 | 0 | 0 |
| Self-perceived COVID-19 recovery | Not recovered | 797 (23.9%) | < 5 | 13 (3.4%) | 35 (9.4%) | 62 (30.1%) | 22 (8.6%) | 111 (18.8%) | 175 (44.1%) | 378 (76.4%) |
| Self-perceived COVID-19 recovery | Recovered | 977 (29.3%) | 62 (9.7%) | 47 (12.4%) | 70 (18.8%) | 29 (14.1%) | 169 (66.0%) | 362 (61.5%) | 161 (40.6%) | 77 (15.6%) |
| Self-perceived COVID-19 recovery | Unknown | 282 (8.5%) | 0 | 0 | 0 | 0 | 65 (25.4%) | 116 (19.7%) | 61 (15.4%) | 40 (8.1%) |
| Weeks between cognitive assessment and symptom start/test date | | 42.0 (29.0, 62.0) | 18.0 (5.0, 39.0) | 20.0 (13.5, 57.0) | 56.0 (35.75, 62.0) | 41.0 (29.0, 59.0) | 53.0 (45.0, 57.25) | 63.0 (40.0, 67.0) | 60.0 (37.0, 67.0) | 38.0 (31.0, 63.0) |

| | | | | | | | | | | |
|---|---|---|---|---|---|---|---|---|---|---|
| Symptom start date/Test date | Q1 Jan-Mar 2020 | 385 (11.5%) | 0 | 8 (2.1%) | 25 (6.7%) | 12 (5.8%) | 0 | 178 (30.2%) | 97 (24.4%) | 65 (13.1%) |
| Symptom start date/Test date | Q2 Apr-Jun 2020 | 879 (26.4%) | 53 (8.3%) | 100 (26.4%) | 168 (45.2%) | 53 (25.7%) | 114 (44.5%) | 186 (31.6%) | 119 (30.0%) | 86 (17.4%) |
| Symptom start date/Test date | Q3 Jul-Sep 2020 | 469 (14.1%) | 92 (14.4%) | 34 (9.0%) | 53 (14.2%) | 35 (17.0%) | 100 (39.1%) | 52 (8.8%) | 55 (13.9%) | 48 (9.7%) |
| Symptom start date/Test date | Q4 Oct-Dec 2020 | 830 (24.9%) | 86 (13.4%) | 27 (7.1%) | 76 (20.4%) | 56 (27.2%) | 40 (15.6%) | 147 (25.0%) | 112 (28.2%) | 286 (57.8%) |
| Symptom start date/Test date | Q1 Jan-Mar 2021 | 319 (9.6%) | 110 (17.2%) | 102 (26.9%) | 36 (9.7%) | 33 (16.0%) | 0 | 17 (2.9%) | 12 (3.0%) | 9 (1.8%) |
| Symptom start date/Test date | Q2 Apr-Jun 2021 | 433 (13.0%) | 296 (46.2%) | 101 (26.6%) | 12 (3.2%) | 16 (7.8%) | < 5 | 5 (0.8%) | < 5 | 0 |
| Symptom start date/Test date | Q3 Jul-Sep 2021 | 20 (0.6%) | < 5 | 7 (1.8%) | < 5 | < 5 | < 5 | < 5 | 0 | < 5 |
| Symptom start date/Test date | Q4 Oct-Dec 2021 | 0 | 0 | 0 | 0 | 0 | 0 | 0 | 0 | 0 |
| Symptom start date/Test date | Q1 Jan-Mar 2022 | 0 | 0 | 0 | 0 | 0 | 0 | 0 | 0 | 0 |
| Symptom start date/Test date | Q2 Apr-Jun 2022 | 0 | 0 | 0 | 0 | 0 | 0 | 0 | 0 | 0 |
| PHQ-4 Scale | Data not available | 36 (1.1%) | < 5 | < 5 | < 5 | < 5 | 5 (2.0%) | 8 (1.4%) | 7 (1.8%) | 6 (1.2%) |
| PHQ-4 Scale | Below threshold, 0-2 (reference) | 2038 (61.8%) | 492 (77.0%) | 227 (60.4%) | 202 (54.6%) | 102 (50.2%) | 197 (78.5%) | 396 (68.2%) | 205 (52.6%) | 217 (44.4%) |
| PHQ-4 Scale | Mild, 3-5 | 896 (27.2%) | 110 (17.2%) | 120 (31.9%) | 103 (27.8%) | 61 (30.0%) | 42 (16.7%) | 145 (25.0%) | 132 (33.8%) | 183 (37.4%) |
| PHQ-4 Scale | Moderate, 6-8 | 258 (7.8%) | 29 (4.5%) | 18 (4.8%) | 45 (12.2%) | 33 (16.3%) | 8 (3.2%) | 32 (5.5%) | 38 (9.7%) | 55 (11.2%) |
| PHQ-4 Scale | Severe, 9-12 | 107 (3.2%) | 8 (1.3%) | 11 (2.9%) | 20 (5.4%) | 7 (3.4%) | < 5 | 8 (1.4%) | 15 (3.8%) | 34 (7.0%) |
| Chalder Fatigue Scale | Data not available | 43 (1.3%) | < 5 | < 5 | < 5 | < 5 | 6 (2.3%) | 10 (1.7%) | 8 (2.0%) | 7 (1.4%) |
| Chalder Fatigue Scale | Below threshold, 0-28 (reference) | 3115 (94.6%) | 634 (99.4%) | 372 (98.9%) | 356 (96.2%) | 181 (89.6%) | 246 (98.4%) | 561 (96.9%) | 359 (92.3%) | 406 (83.2%) |
| Chalder Fatigue Scale | Above threshold, 29-33 | 177 (5.4%) | < 5 | < 5 | 14 (3.8%) | 21 (10.4%) | < 5 | 18 (3.1%) | 30 (7.7%) | 82 (16.8%) |
| WSAS | Data not available | 50 (1.5%) | 5 (0.8%) | < 5 | < 5 | < 5 | 9 (3.5%) | 10 (1.7%) | 9 (2.3%) | 7 (1.4%) |
| WSAS | Below threshold, 0-9 (reference) | 2227 (67.8%) | 550 (86.5%) | 269 (71.5%) | 226 (61.2%) | 93 (46.0%) | 214 (86.6%) | 455 (78.6%) | 225 (58.0%) | 195 (40.0%) |
| WSAS | Mild, 10-20 | 714 (21.7%) | 67 (10.5%) | 80 (21.3%) | 106 (28.7%) | 55 (27.2%) | 24 (9.7%) | 96 (16.6%) | 124 (32.0%) | 162 (33.2%) |
| WSAS | Moderate to severe, 21-40 | 344 (10.5%) | 19 (3.0%) | 27 (7.2%) | 37 (10.0%) | 54 (26.7%) | 9 (3.6%) | 28 (4.8%) | 39 (10.1%) | 131 (26.8%) |

#### S8. Factors associated with participation

Evidence of association with participation in cognitive testing in both rounds of cognitive testing was found for the following groups:

1. Individuals with a higher number of non-responses to previous invitations to CSSB studies were less likely to participate.
2. Older age groups were more likely to participate than younger age groups.
3. Participants who joined the CSSB in the later second round of recruitment in May 2021 were more likely to participate than the first recruitment cohort who joined in Oct-Nov 2020.
4. After controlling for COVID-19 group at time of recruitment (isolating the effect of change in group between recruitment and invitation), those with prior illness of ≥ 12 weeks symptom duration at time of invitation to cognitive assessment were more likely to participate with the SARS-CoV-2 positive group.
5. Male sex individuals were less likely to participate than female sex

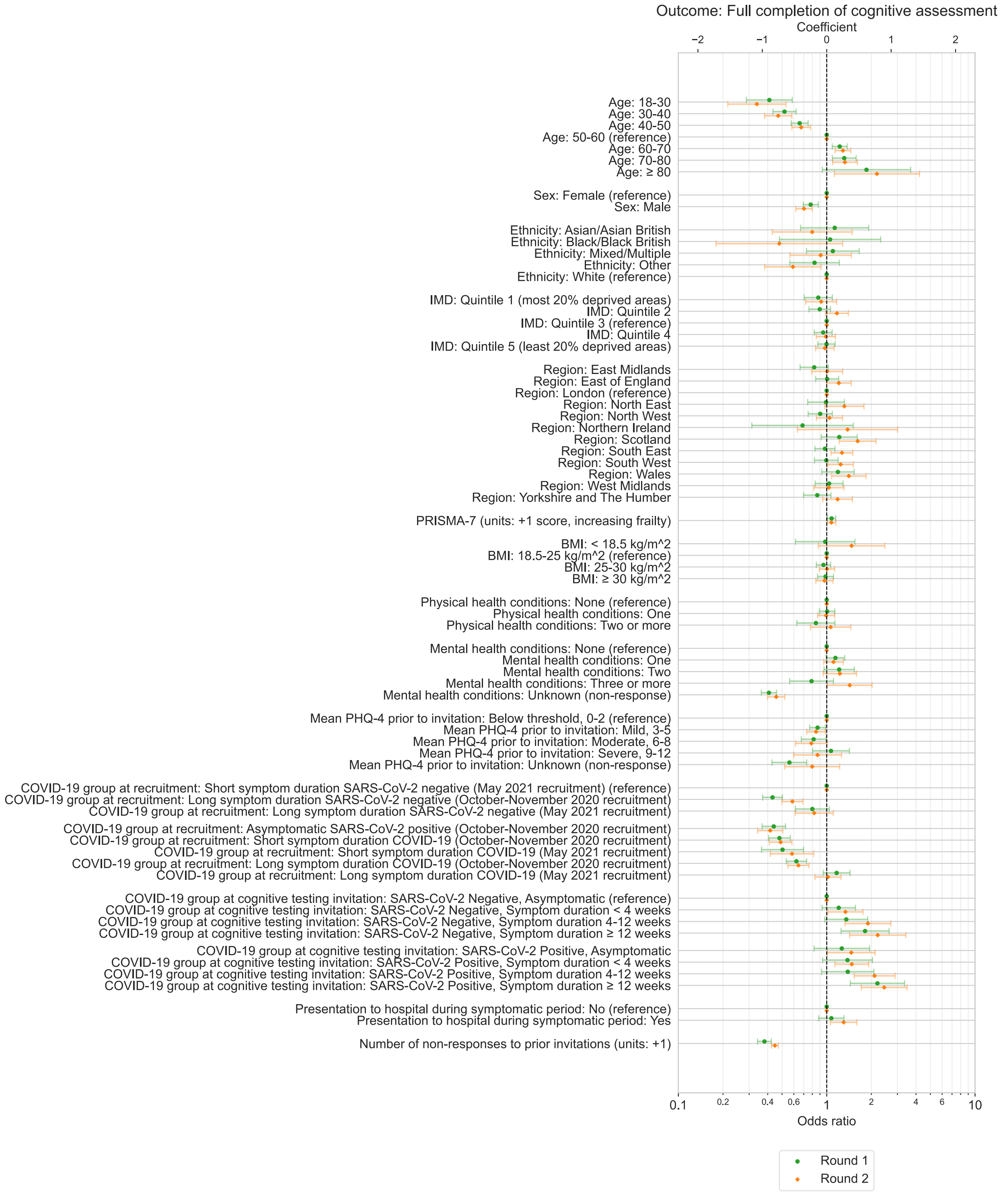


Figure S 3. **Associations with participation in individual rounds of cognitive assessment**. Odds ratios with 95% confidence intervals from multivariable logistic regression models, testing association between exposure variables and full completion following invitation to either Round 1 or Round 2 of cognitive testing. Results for each exposure variable presented originate from separate models that use distinct adjustment variable sets determined from the proposed DAG for participation.

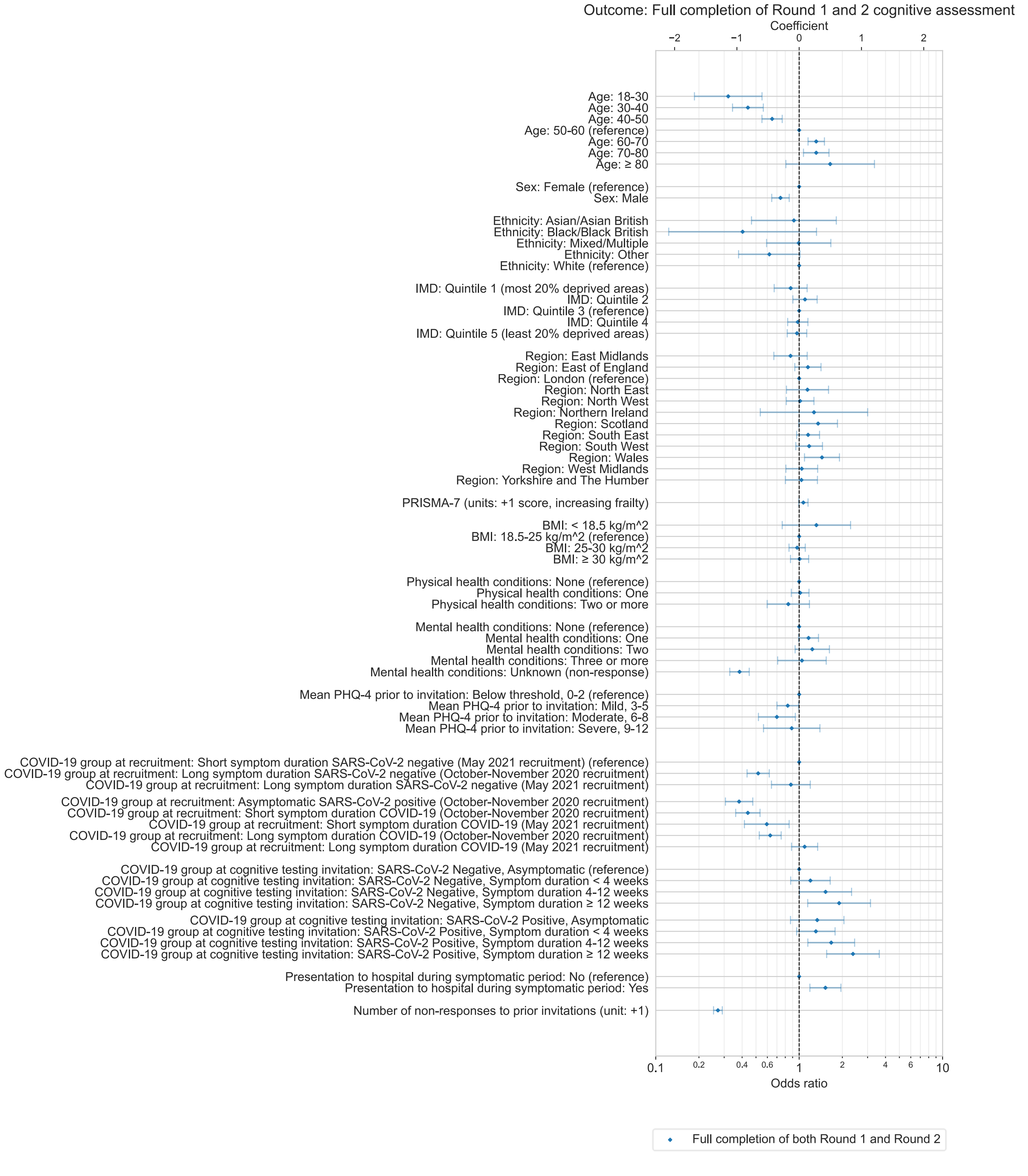


Figure S 4. **Associations with participation in both rounds of cognitive assessment**. Odds ratios with 95% confidence intervals from multivariable logistic regression models, testing association between exposure variables and full completion of both Round 1 and Round 2 of cognitive testing following invitation. Results for each exposure variable presented originate from separate models that use distinct adjustment variable sets determined from the proposed DAG for participation.

Table S 4. **Associations with participation in cognitive testing.** Odds ratios with 95% confidence intervals from multivariable logistic regression models, testing association between exposure variables and full completion of Round 1 and/or Round 2 of cognitive testing following invitation. Results for each exposure variable presented originate from separate models that use distinct adjustment variable sets determined from the proposed DAG for participation.

| | Round 1 participation (full) | | | Round 2 participation (full) | | | Round 1 and Round 2 participation (full) | | |
|---|---|---|---|---|---|---|---|---|---|
| Exposure variable | OR | Lower 95% CI | Upper 95% CI | OR | Lower 95% CI | Upper 95% CI | OR | Lower 95% CI | Upper 95% CI |
| Age: 18-30 | 0.41 | 0.29 | 0.59 | 0.34 | 0.22 | 0.53 | 0.32 | 0.19 | 0.55 |
| Age: 30-40 | 0.52 | 0.43 | 0.62 | 0.47 | 0.38 | 0.58 | 0.44 | 0.34 | 0.56 |
| Age: 40-50 | 0.66 | 0.58 | 0.75 | 0.67 | 0.58 | 0.78 | 0.65 | 0.55 | 0.76 |
| Age: 50-60 (reference) | 1.00 | | | 1.00 | | | 1.00 | | |
| Age: 60-70 | 1.22 | 1.09 | 1.38 | 1.29 | 1.14 | 1.45 | 1.31 | 1.15 | 1.50 |
| Age: 70-80 | 1.31 | 1.09 | 1.58 | 1.33 | 1.10 | 1.61 | 1.31 | 1.07 | 1.61 |
| Age: ≥ 80 | 1.85 | 0.93 | 3.68 | 2.18 | 1.13 | 4.22 | 1.64 | 0.81 | 3.35 |
| Sex: Female (reference) | 1.00 | | | 1.00 | | | 1.00 | | |
| Sex: Male | 0.78 | 0.69 | 0.88 | 0.70 | 0.62 | 0.80 | 0.74 | 0.64 | 0.85 |
| Ethnicity: Asian/Asian British | 1.13 | 0.67 | 1.92 | 0.80 | 0.43 | 1.48 | 0.92 | 0.46 | 1.81 |
| Ethnicity: Black/Black British | 1.05 | 0.48 | 2.31 | 0.48 | 0.18 | 1.28 | 0.40 | 0.12 | 1.32 |
| Ethnicity: Mixed/Multiple | 1.10 | 0.73 | 1.66 | 0.91 | 0.57 | 1.47 | 0.99 | 0.59 | 1.66 |
| Ethnicity: Other | 0.83 | 0.56 | 1.22 | 0.59 | 0.38 | 0.92 | 0.62 | 0.38 | 1.02 |
| Ethnicity: White (reference) | 1.00 | | | 1.00 | | | 1.00 | | |
| IMD: Quintile 1 (most 20% deprived areas) | 0.88 | 0.70 | 1.09 | 0.92 | 0.72 | 1.17 | 0.87 | 0.67 | 1.14 |
| IMD: Quintile 2 | 0.90 | 0.76 | 1.06 | 1.17 | 0.98 | 1.40 | 1.10 | 0.90 | 1.33 |
| IMD: Quintile 3 (reference) | 1.00 | | | 1.00 | | | 1.00 | | |
| IMD: Quintile 4 | 0.95 | 0.82 | 1.09 | 0.99 | 0.85 | 1.15 | 0.98 | 0.83 | 1.15 |
| IMD: Quintile 5 (least 20% deprived areas) | 1.00 | 0.87 | 1.14 | 0.97 | 0.84 | 1.12 | 0.97 | 0.83 | 1.13 |
| Region: East Midlands | 0.82 | 0.66 | 1.03 | 1.01 | 0.79 | 1.28 | 0.87 | 0.67 | 1.14 |
| Region: East of England | 1.01 | 0.84 | 1.20 | 1.21 | 0.99 | 1.46 | 1.15 | 0.93 | 1.42 |
| Region: London (reference) | 1.00 | | | 1.00 | | | 1.00 | | |
| Region: North East | 0.99 | 0.74 | 1.32 | 1.32 | 0.97 | 1.78 | 1.14 | 0.82 | 1.60 |
| Region: North West | 0.90 | 0.75 | 1.09 | 1.04 | 0.85 | 1.28 | 1.01 | 0.81 | 1.27 |
| Region: Northern Ireland | 0.69 | 0.31 | 1.51 | 1.38 | 0.63 | 3.01 | 1.27 | 0.54 | 3.00 |

| | | | | | | | | | |
|---|---|---|---|---|---|---|---|---|---|
| Region: Scotland | 1.21 | 0.92 | 1.61 | 1.62 | 1.22 | 2.15 | 1.35 | 0.99 | 1.85 |
| Region: South East | 0.97 | 0.83 | 1.14 | 1.27 | 1.07 | 1.50 | 1.15 | 0.96 | 1.39 |
| Region: South West | 0.99 | 0.83 | 1.19 | 1.24 | 1.02 | 1.51 | 1.17 | 0.95 | 1.45 |
| Region: Wales | 1.19 | 0.93 | 1.53 | 1.41 | 1.08 | 1.84 | 1.44 | 1.09 | 1.91 |
| Region: West Midlands | 1.04 | 0.84 | 1.29 | 1.03 | 0.82 | 1.31 | 1.04 | 0.81 | 1.35 |
| Region: Yorkshire and The Humber | 0.86 | 0.70 | 1.07 | 1.18 | 0.94 | 1.49 | 1.04 | 0.80 | 1.34 |
| PRISMA-7 (units: +1 score, increasing frailty) | 1.08 | 1.01 | 1.15 | 1.08 | 1.00 | 1.15 | 1.07 | 0.99 | 1.15 |
| BMI: < 18.5 kg/m^2 | 0.98 | 0.62 | 1.55 | 1.47 | 0.88 | 2.47 | 1.32 | 0.76 | 2.28 |
| BMI: 18.5-25 kg/m^2 (reference) | 1.00 | | | 1.00 | | | 1.00 | | |
| BMI: 25-30 kg/m^2 | 0.95 | 0.85 | 1.06 | 1.00 | 0.89 | 1.13 | 0.97 | 0.85 | 1.10 |
| BMI: ≥ 30 kg/m^2 | 0.98 | 0.87 | 1.11 | 0.97 | 0.84 | 1.10 | 1.01 | 0.87 | 1.16 |
| Physical health conditions: None (reference) | 1.00 | | | 1.00 | | | 1.00 | | |
| Physical health conditions: One | 1.01 | 0.89 | 1.14 | 0.99 | 0.87 | 1.13 | 1.02 | 0.88 | 1.17 |
| Physical health conditions: Two or more | 0.85 | 0.63 | 1.14 | 1.06 | 0.78 | 1.46 | 0.84 | 0.60 | 1.18 |
| Mental health conditions: None (reference) | 1.00 | | | 1.00 | | | 1.00 | | |
| Mental health conditions: One | 1.15 | 0.99 | 1.32 | 1.11 | 0.96 | 1.29 | 1.16 | 0.99 | 1.36 |
| Mental health conditions: Two | 1.21 | 0.96 | 1.54 | 1.23 | 0.95 | 1.59 | 1.23 | 0.94 | 1.63 |
| Mental health conditions: Three or more | 0.79 | 0.56 | 1.11 | 1.43 | 1.01 | 2.02 | 1.05 | 0.71 | 1.54 |
| Mental health conditions: Unknown (non-response) | 0.41 | 0.36 | 0.46 | 0.46 | 0.40 | 0.52 | 0.38 | 0.33 | 0.45 |
| Mean PHQ-4 prior to invitation: Below threshold, 0-2 (reference) | 1.00 | | | 1.00 | | | 1.00 | | |
| Mean PHQ-4 prior to invitation: Mild, 3-5 | 0.87 | 0.77 | 0.98 | 0.85 | 0.73 | 0.98 | 0.83 | 0.70 | 0.99 |
| Mean PHQ-4 prior to invitation: Moderate, 6-8 | 0.82 | 0.67 | 0.99 | 0.79 | 0.62 | 1.01 | 0.70 | 0.52 | 0.94 |
| Mean PHQ-4 prior to invitation: Severe, 9-12 | 1.07 | 0.80 | 1.42 | 0.87 | 0.60 | 1.26 | 0.89 | 0.56 | 1.40 |
| Mean PHQ-4 prior to invitation: Unknown (non-response) | 0.56 | 0.43 | 0.73 | 0.80 | 0.52 | 1.22 | | | |
| COVID-19 group at recruitment: Short symptom duration SARS-CoV-2 negative (May 2021 recruitment) (reference) | 1.00 | | | 1.00 | | | 1.00 | | |
| COVID-19 group at recruitment: Long symptom duration SARS-CoV-2 negative (October-November 2020 recruitment) | 0.43 | 0.37 | 0.50 | 0.59 | 0.50 | 0.69 | 0.52 | 0.43 | 0.62 |
| COVID-19 group at recruitment: Long symptom duration SARS-CoV-2 negative (May 2021 recruitment) | 0.80 | 0.62 | 1.04 | 0.82 | 0.61 | 1.11 | 0.88 | 0.64 | 1.19 |
| COVID-19 group at recruitment: Asymptomatic SARS-CoV-2 positive (October-November 2020 recruitment) | 0.44 | 0.37 | 0.53 | 0.42 | 0.34 | 0.51 | 0.38 | 0.31 | 0.47 |

| | | | | | | | | | |
|---|---|---|---|---|---|---|---|---|---|
| COVID-19 group at recruitment: Short symptom duration COVID-19 (October-November 2020 recruitment) | 0.48 | 0.41 | 0.57 | 0.49 | 0.41 | 0.58 | 0.44 | 0.36 | 0.53 |
| COVID-19 group at recruitment: Short symptom duration COVID-19 (May 2021 recruitment) | 0.50 | 0.36 | 0.70 | 0.58 | 0.42 | 0.82 | 0.59 | 0.42 | 0.85 |
| COVID-19 group at recruitment: Long symptom duration COVID-19 (October-November 2020 recruitment) | 0.62 | 0.53 | 0.73 | 0.64 | 0.55 | 0.76 | 0.63 | 0.53 | 0.75 |
| COVID-19 group at recruitment: Long symptom duration COVID-19 (May 2021 recruitment) | 1.17 | 0.95 | 1.44 | 1.02 | 0.83 | 1.25 | 1.09 | 0.88 | 1.35 |
| COVID-19 group at cognitive testing invitation: SARS-CoV-2 Negative, Asymptomatic (reference) | 1.00 | | | 1.00 | | | 1.00 | | |
| COVID-19 group at cognitive testing invitation: SARS-CoV-2 Negative, Symptom duration < 4 weeks | 1.20 | 0.93 | 1.56 | 1.34 | 1.02 | 1.76 | 1.20 | 0.87 | 1.65 |
| COVID-19 group at cognitive testing invitation: SARS-CoV-2 Negative, Symptom duration 4-12 weeks | 1.35 | 0.97 | 1.89 | 1.90 | 1.33 | 2.71 | 1.53 | 1.01 | 2.32 |
| COVID-19 group at cognitive testing invitation: SARS-CoV-2 Negative, Symptom duration ≥ 12 weeks | 1.81 | 1.25 | 2.63 | 2.21 | 1.43 | 3.42 | 1.90 | 1.15 | 3.14 |
| COVID-19 group at cognitive testing invitation: SARS-CoV-2 Positive, Asymptomatic | 1.26 | 0.82 | 1.95 | 1.46 | 1.01 | 2.12 | 1.34 | 0.87 | 2.05 |
| COVID-19 group at cognitive testing invitation: SARS-CoV-2 Positive, Symptom duration < 4 weeks | 1.38 | 0.94 | 2.03 | 1.48 | 1.14 | 1.92 | 1.31 | 0.96 | 1.78 |
| COVID-19 group at cognitive testing invitation: SARS-CoV-2 Positive, Symptom duration 4-12 weeks | 1.39 | 0.92 | 2.08 | 2.10 | 1.53 | 2.89 | 1.67 | 1.15 | 2.44 |
| COVID-19 group at cognitive testing invitation: SARS-CoV-2 Positive, Symptom duration ≥ 12 weeks | 2.20 | 1.44 | 3.35 | 2.44 | 1.71 | 3.49 | 2.37 | 1.56 | 3.62 |
| Presentation to hospital during symptomatic period: No (reference) | 1.00 | | | 1.00 | | | 1.00 | | |
| Presentation to hospital during symptomatic period: Yes | 1.07 | 0.88 | 1.31 | 1.30 | 1.06 | 1.60 | 1.52 | 1.19 | 1.96 |
| Number of non-responses to prior invitations (units: +1) | 0.38 | 0.34 | 0.42 | 0.45 | 0.42 | 0.47 | 0.27 | 0.25 | 0.29 |

#### S9. Predictive modelling for inverse probability weight generation

Table S 5. **Summary of logistic regression models used to generate inverse probability of response weights.**

| | Highest performing model | | Weight-generating model | | Scaled weight (median, IQR) | |
|---|---|---|---|---|---|---|
| Model | Variable set | AUC-ROC | Variable set | AUC-ROC | Did not participate | Participated |
| Round 1 full completion | Prior non-response count, COVID-19 group at invitation, Age, COVID-19 group at recruitment, PHQ-4 category, Mental health condition count, Physical health condition count, PRISMA-7 score | 0.727 | Age, Sex, Ethnicity, Deprivation, Region, BMI, Physical health condition count, Mental health condition count, PHQ-4 category, PRISMA-7 score, COVID-19 group at recruitment, COVID-19 group at invitation, Presentation to hospital, Prior non-response count | 0.719 | 0.71 (0.51, 1.36) | 0.49 (0.41, 0.61) |
| Round 2 full completion | Prior non-response count, Age, Mental health condition count, Sex, Deprivation, Presentation to hospital, PRISMA-7 score, PHQ-4 category, Ethnicity, Physical health condition count | 0.825 | | 0.820 | 0.68 (0.31, 1.60) | 0.21 (0.17, 0.32) |
| Round 1 and 2 full completion | Prior non-response count, Presentation to hospital, Mental health condition count, Sex, Age, Deprivation, Physical health condition count, PRISMA-7 score | 0.874 | | 0.872 | 0.29 (0.07, 1.23) | 0.03 (0.02, 0.05) |

#### S10. Principal component analysis

The first principal components represent higher task accuracy, larger average reaction time, or larger reaction time variation across all tasks, while the second principal components represented variation due to opposing trends between certain tasks. For task accuracy, the second principal component primarily represents variation due to poorer performance in the four immediate and delayed memory tasks and other tasks. For average reaction time or reaction time variation, the second principal component represents variation due to shorter (relative to other tasks) average reaction time or lower variation in the following tasks: immediate and delayed word-based memory tasks, motor control, cognitive reflection test and spatial span.

Table S 6. **Principal component analysis of Round 1 cognitive assessment metrics**. Explained variance, eigenvalues and loadings from individual cognitive assessment tasks for components obtained from principal component analysis of Round 1 cognitive assessment accuracy, average reaction time and within-task reaction time variation metrics for factors with eigenvalues greater or equal to one.

| | **Accuracy** | | | **Average reaction time** | | **Reaction time variation** | | |
|---|---|---|---|---|---|---|---|---|
| **Task** | **PCA component 1** | **PCA component 2** | **PCA component 3** | **PCA component 1** | **PCA component 2** | **PCA component 1** | **PCA component 2** | **PCA component 3** |
| Immediate memory (words) | 0.30 | -0.22 | -0.30 | 0.36 | -0.20 | 0.34 | -0.39 | 0.22 |
| Immediate memory (objects) | 0.40 | -0.39 | -0.01 | 0.27 | 0.42 | 0.33 | 0.30 | 0.01 |
| Motor control | 0.14 | -0.15 | 0.81 | 0.31 | -0.40 | 0.23 | -0.37 | 0.13 |
| 2-D mental manipulations | 0.28 | 0.40 | -0.15 | 0.23 | 0.35 | 0.31 | 0.24 | 0.14 |
| Target detection | 0.21 | 0.33 | -0.09 | 0.23 | -0.14 | 0.07 | 0.32 | 0.70 |
| Spatial span | 0.28 | 0.39 | -0.01 | 0.27 | -0.26 | 0.23 | -0.11 | -0.31 |
| Tower of London | 0.29 | 0.22 | 0.17 | 0.19 | 0.24 | 0.21 | 0.15 | -0.45 |
| Verbal analogies | 0.31 | 0.24 | 0.09 | 0.25 | 0.31 | 0.33 | 0.22 | -0.09 |
| Delayed memory (words) | 0.29 | -0.25 | -0.26 | 0.36 | -0.12 | 0.38 | -0.22 | 0.16 |
| Delayed memory (objects) | 0.40 | -0.40 | -0.01 | 0.30 | 0.39 | 0.35 | 0.33 | 0.05 |
| Paired-associate learning | 0.28 | 0.14 | -0.04 | 0.34 | -0.02 | 0.28 | 0.13 | -0.31 |
| Cognitive reflection test | 0.18 | 0.05 | 0.34 | 0.29 | -0.31 | 0.27 | -0.45 | 0.03 |
| *Variance explained* | 26% | 12% | 10% | 44% | 12% | 25% | 13% | 9% |
| *Eigenvalue* | 3.13 | 1.43 | 1.21 | 5.35 | 1.50 | 2.92 | 1.51 | 1.08 |

Table S 7. **Principal component analysis of Round 2 cognitive assessment metrics**. Explained variance, eigenvalues and loadings from individual cognitive assessment tasks for components obtained from principal component analysis of Round 2 cognitive assessment accuracy, average reaction time and within-task reaction time variation metrics for factors with eigenvalues greater or equal to one.

| | **Accuracy** | | | **Average reaction time** | | **Reaction time variation** | | |
|---|---|---|---|---|---|---|---|---|
| **Task** | **PCA component 1** | **PCA component 2** | **PCA component 3** | **PCA component 1** | **PCA component 2** | **PCA component 1** | **PCA component 2** | **PCA component 3** |
| Immediate memory (words) | 0.28 | -0.28 | -0.20 | 0.37 | 0.16 | 0.36 | 0.35 | 0.12 |
| Immediate memory (objects) | 0.39 | -0.37 | 0.00 | 0.25 | -0.36 | 0.33 | -0.30 | 0.23 |
| Motor control | 0.08 | -0.05 | 0.32 | 0.29 | 0.37 | 0.27 | 0.43 | 0.09 |
| 2-D mental manipulations | 0.30 | 0.42 | -0.22 | 0.24 | -0.37 | 0.26 | -0.28 | 0.21 |
| Target detection | 0.21 | 0.34 | -0.46 | 0.25 | 0.20 | 0.10 | -0.05 | 0.30 |
| Spatial span | 0.25 | 0.37 | 0.07 | 0.27 | 0.28 | 0.23 | 0.19 | -0.57 |
| Tower of London | 0.30 | 0.15 | 0.34 | 0.19 | -0.32 | 0.24 | -0.28 | -0.57 |
| Verbal analogies | 0.32 | 0.28 | 0.18 | 0.27 | -0.36 | 0.32 | -0.26 | -0.02 |
| Delayed memory (words) | 0.32 | -0.28 | -0.19 | 0.37 | 0.11 | 0.38 | 0.20 | 0.13 |
| Delayed memory (objects) | 0.41 | -0.38 | -0.02 | 0.29 | -0.33 | 0.33 | -0.35 | 0.19 |
| Paired-associate learning | 0.28 | 0.15 | 0.00 | 0.32 | 0.04 | 0.25 | -0.13 | -0.30 |
| Cognitive reflection test | 0.13 | 0.02 | 0.64 | 0.31 | 0.30 | 0.29 | 0.40 | 0.02 |
| *Variance explained* | 27% | 12% | 9% | 44% | 13% | 25% | 13% | 9% |
| *Eigenvalue* | 3.12 | 1.41 | 1.04 | 5.00 | 1.49 | 3.01 | 1.52 | 1.07 |

#### S11. Cognitive accuracy – PCA 1st component, all exposures

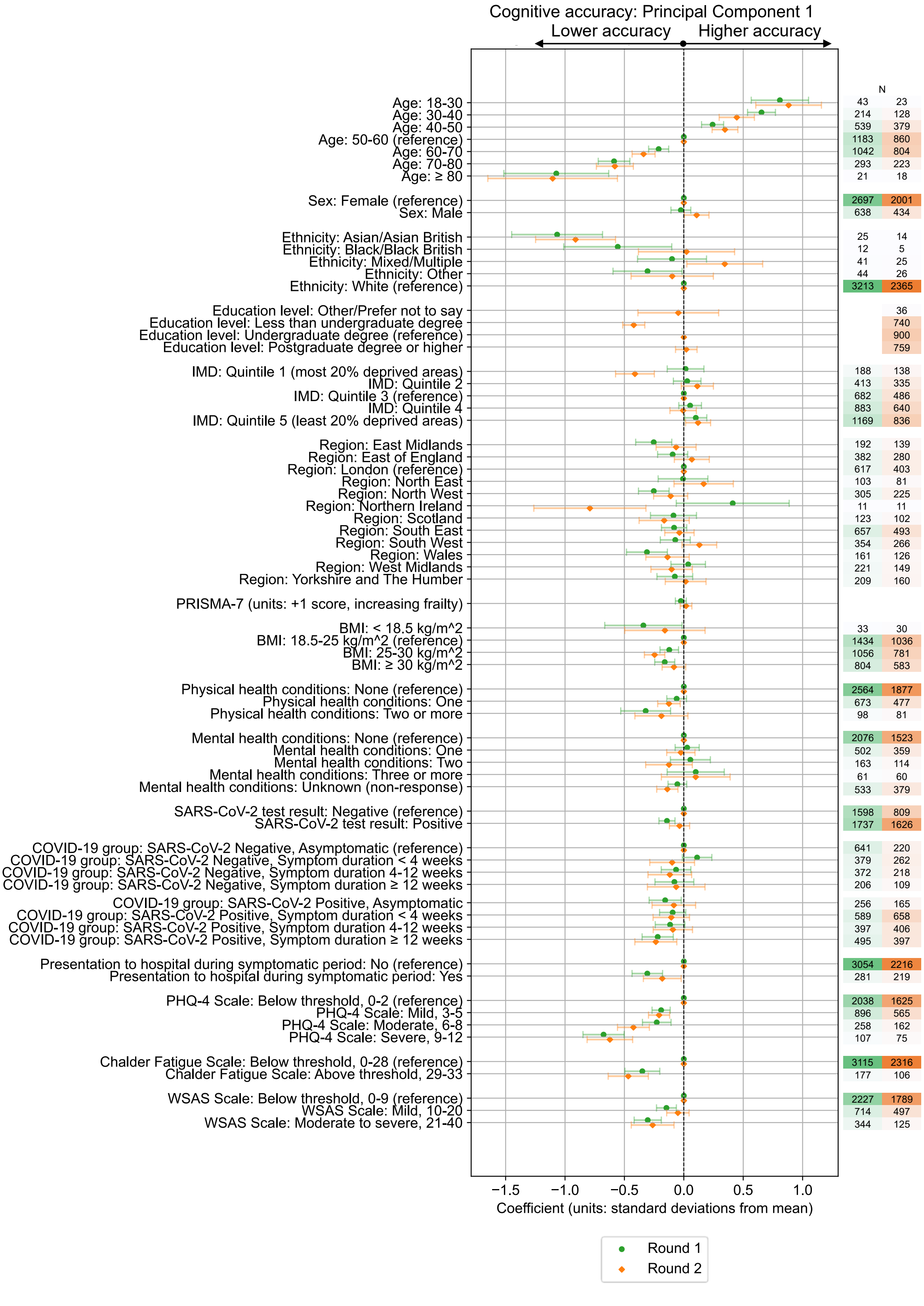


Figure S 5. **Associations with composite cognitive assessment accuracy scores**. Standardised coefficients (number of standard deviations from mean) with 95% confidence intervals from multivariable ordinary least squares linear regression models testing association between various exposure variables and composite cognitive accuracy scores from 1st principal component. Results are from Round 1 and Round 2 of cognitive testing, among all individuals who completed either Round 1 or Round 2 of testing. Results for each exposure variable originate from separate models that use distinct adjustment variable sets determined from the proposed DAG for cognitive performance.

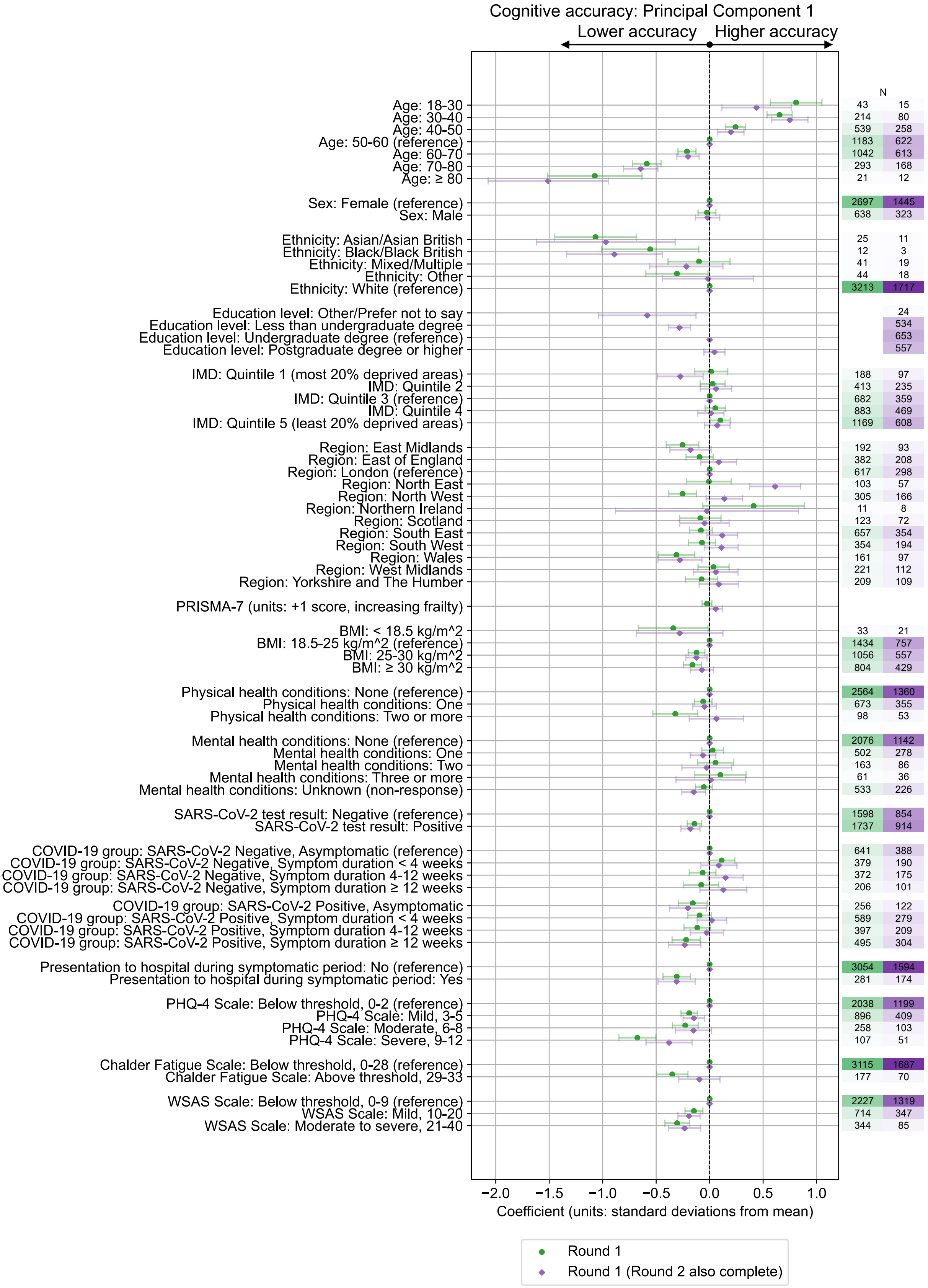


Figure S 6. **Associations with composite cognitive assessment accuracy scores**. Standardised coefficients (number of standard deviations from mean) with 95% confidence intervals from multivariable ordinary least squares linear regression models testing association between various exposure variables and composite cognitive accuracy scores from 1st principal component. Results are from Round 1 of cognitive testing, for all individuals who completed Round 1 (green circle markers), or individuals who completed both Round 1 and Round 2 (purple diamond markers). Results for each exposure variable originate from separate models that use distinct adjustment variable sets determined from the proposed DAG for cognitive performance.

#### S12. Cognitive accuracy – Round 1 association with COVID-19 group by individual task

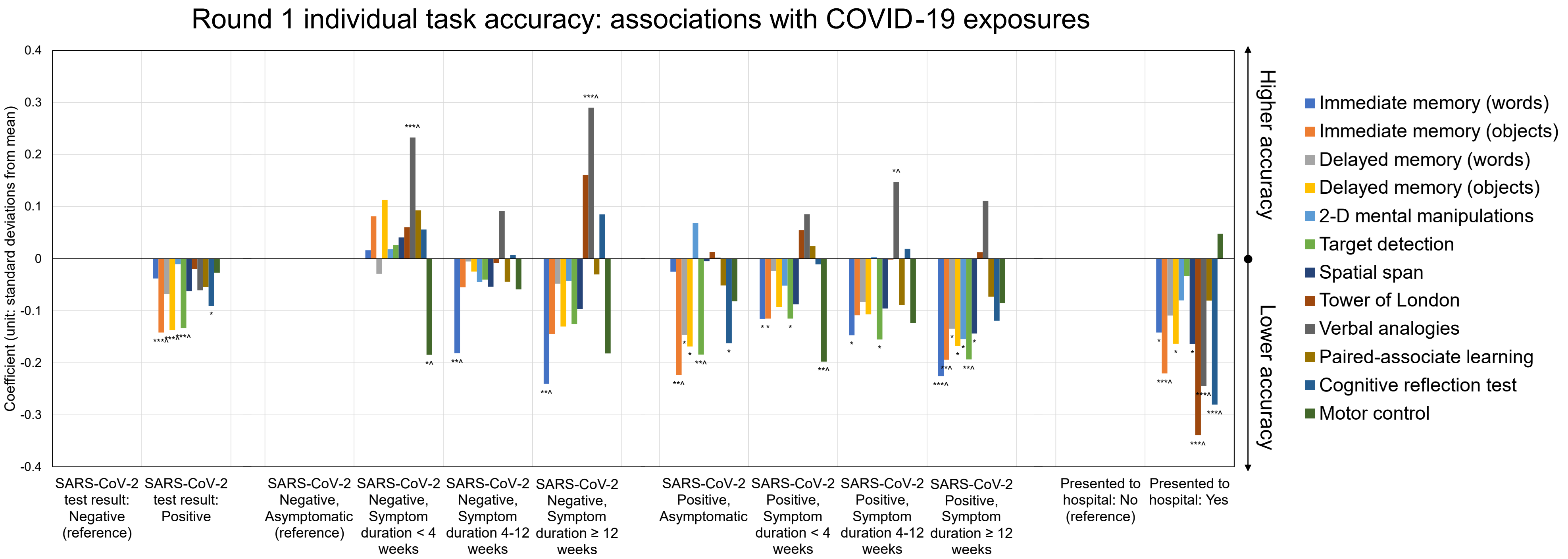


Figure S 7. **Association between COVID-19 related exposures and individual task accuracy scores**. Standardised coefficients (number of standard deviations from mean) with indication of p-values from multivariable ordinary least squares linear regression models testing association between COVID-19 group exposures and individual task accuracy standardised scores. Results are from Round 1 of cognitive testing, among all those who fully completed all tasks. Models used the following set of adjustment variables: Age, BMI, Deprivation, Ethnicity, Frailty (PRISMA-7), Mental health condition count, Physical health condition count, Presentation to hospital, Region, Sex. Asterisk symbols represent uncorrected p-values, * < 0.05, ** < 0.01, *** < 0.001, while caret symbol ^ illustrates p < 0.05 after adjustment for multiple testing.

#### S13. Cognitive accuracy – full heatmaps

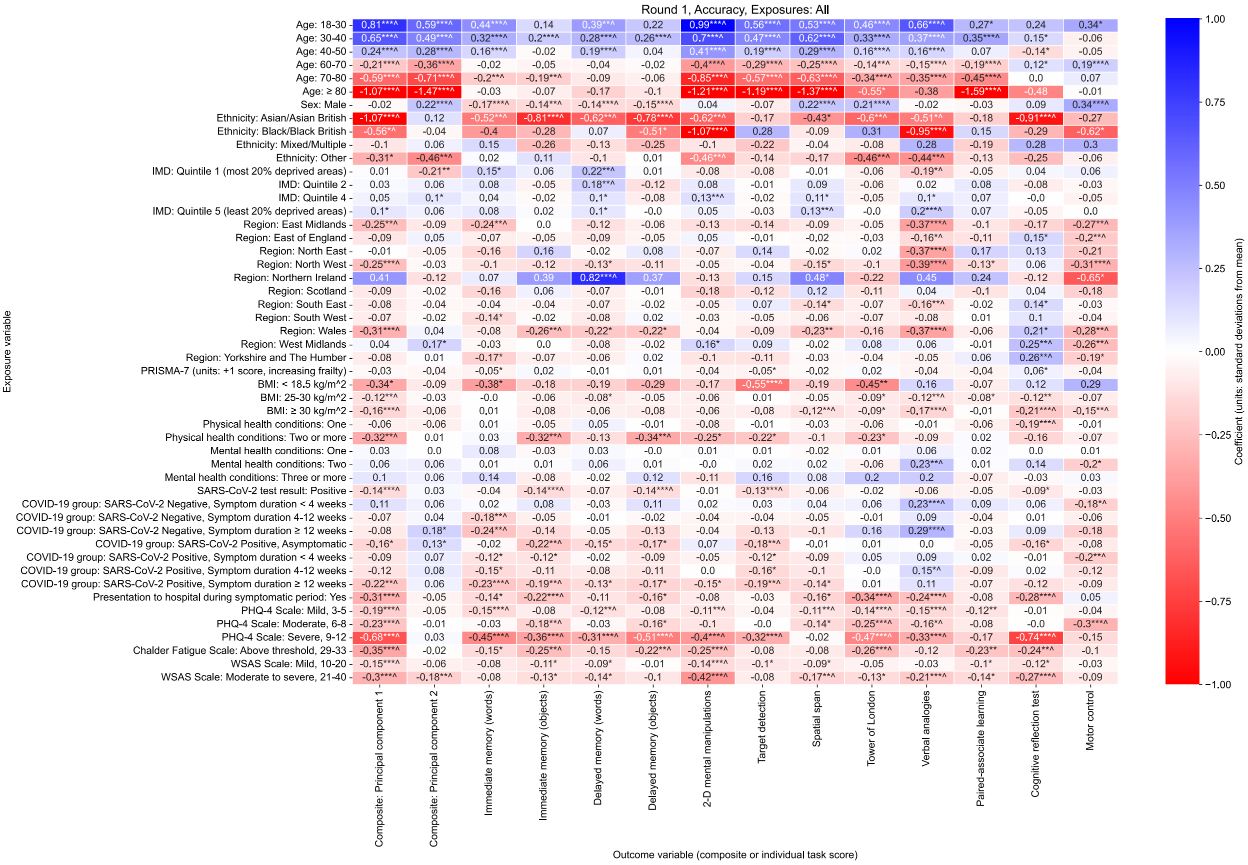


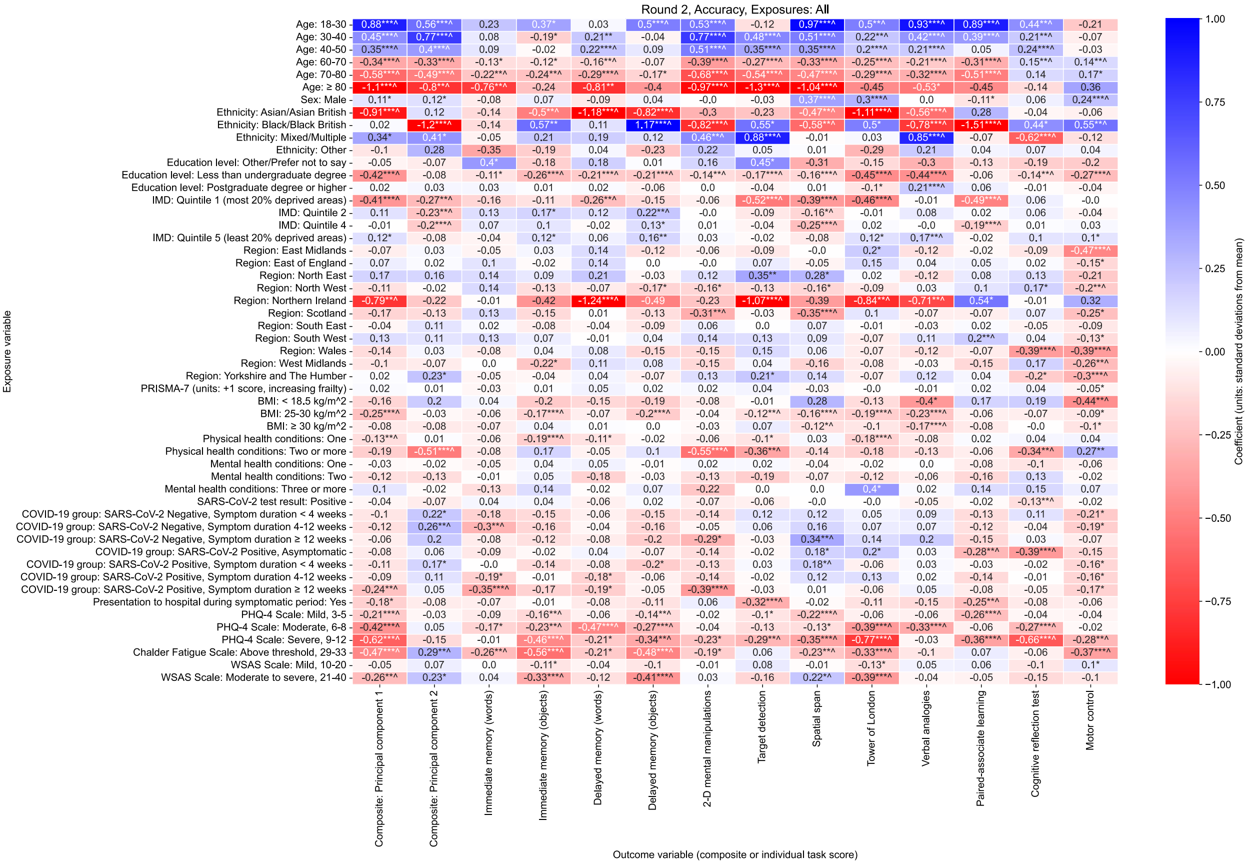


Figure S 8. **Associations with cognitive assessment accuracy scores**. Standardised coefficients (number of standard deviations from mean) from multivariable ordinary least squares linear regression models testing association between various exposure variables and accuracy scores during cognitive tasks as outcome variables. Results are from Round 1 and Round 2 of cognitive testing, among all individuals who completed either Round 1 or Round 2 of testing. Asterisk symbols represent uncorrected p-values, * < 0.05, ** < 0.01, *** < 0.001, while superscript caret symbol ^ illustrates $p < 0.05$ after adjustment for multiple testing. Results for each combination of both exposure variable and outcome variable presented originate from separate models that use distinct adjustment variable sets determined from the proposed DAG for cognitive performance.

### S14. Within-task variation in reaction time heatmaps

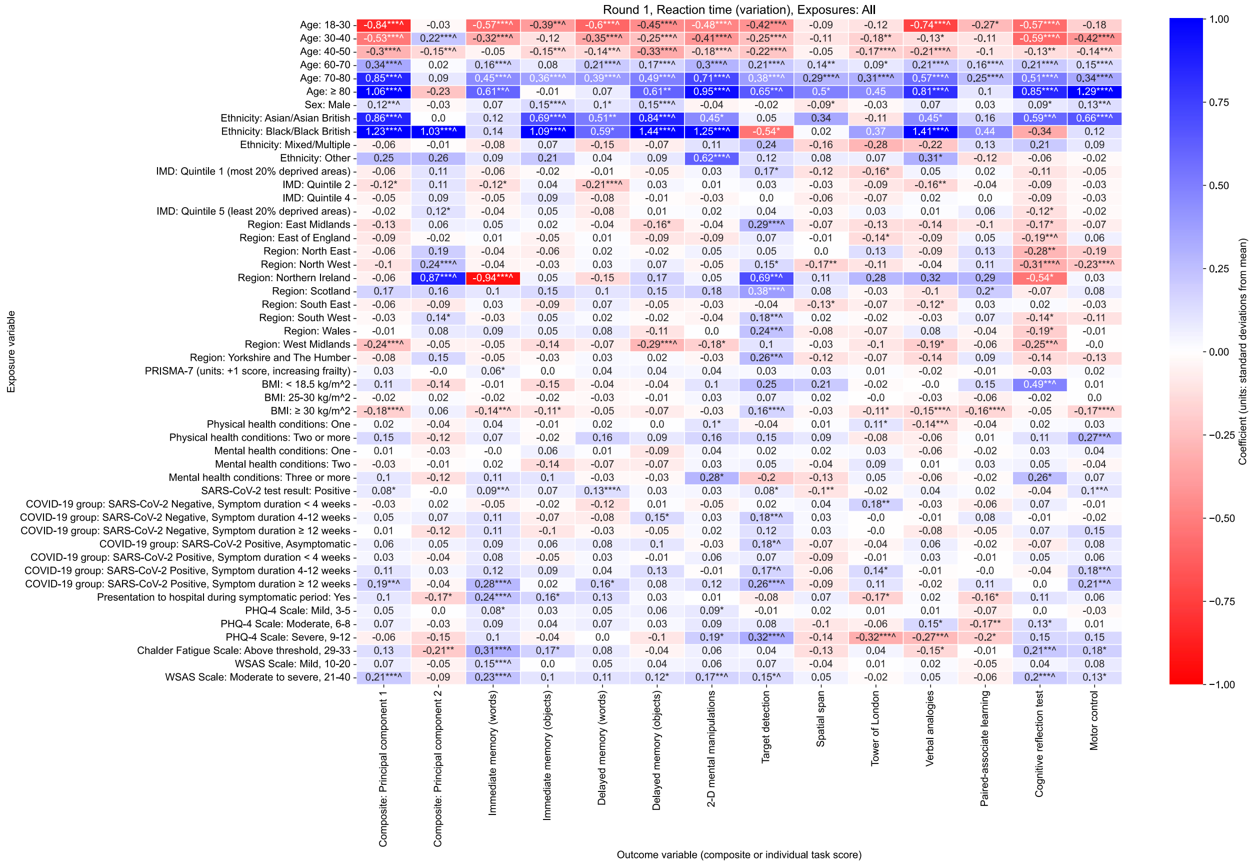


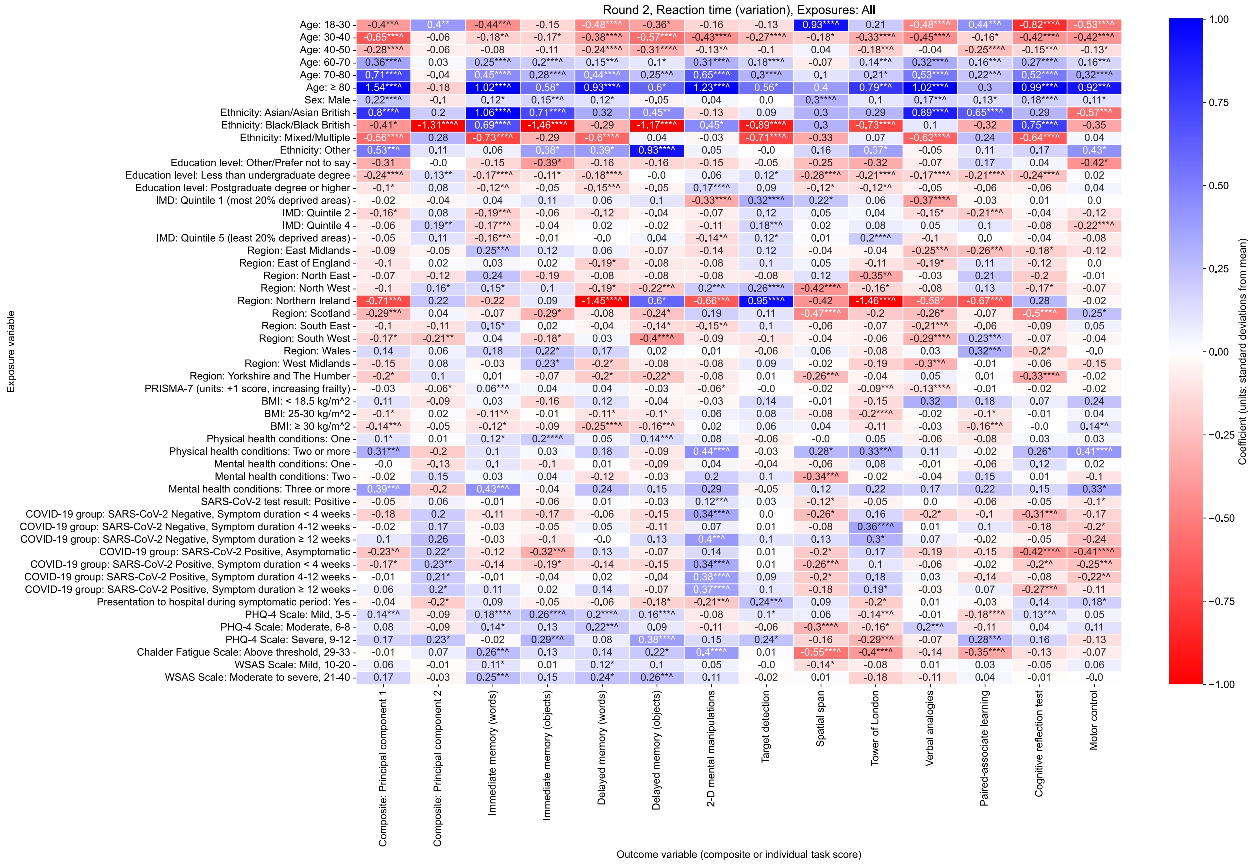


Figure S 9. **Associations with cognitive assessment reaction time variation**. Standardised coefficients (number of standard deviations from mean) from multivariable ordinary least squares linear regression models testing association between various exposure variables and variation in reaction time within cognitive tasks as outcome variables. Results are from Round 1 and Round 2 of cognitive testing, among all individuals who completed Round 1 or Round 2 of testing. Asterisk symbols represent uncorrected p-values, * < 0.05, ** < 0.01, *** < 0.001, while superscript caret symbol ^ illustrates $p < 0.05$ after adjustment for multiple testing. Results for each combination of both exposure variable and outcome variable presented originate from separate models that use distinct adjustment variable sets determined from the proposed DAG for cognitive performance.

### S15. Average reaction time heatmaps

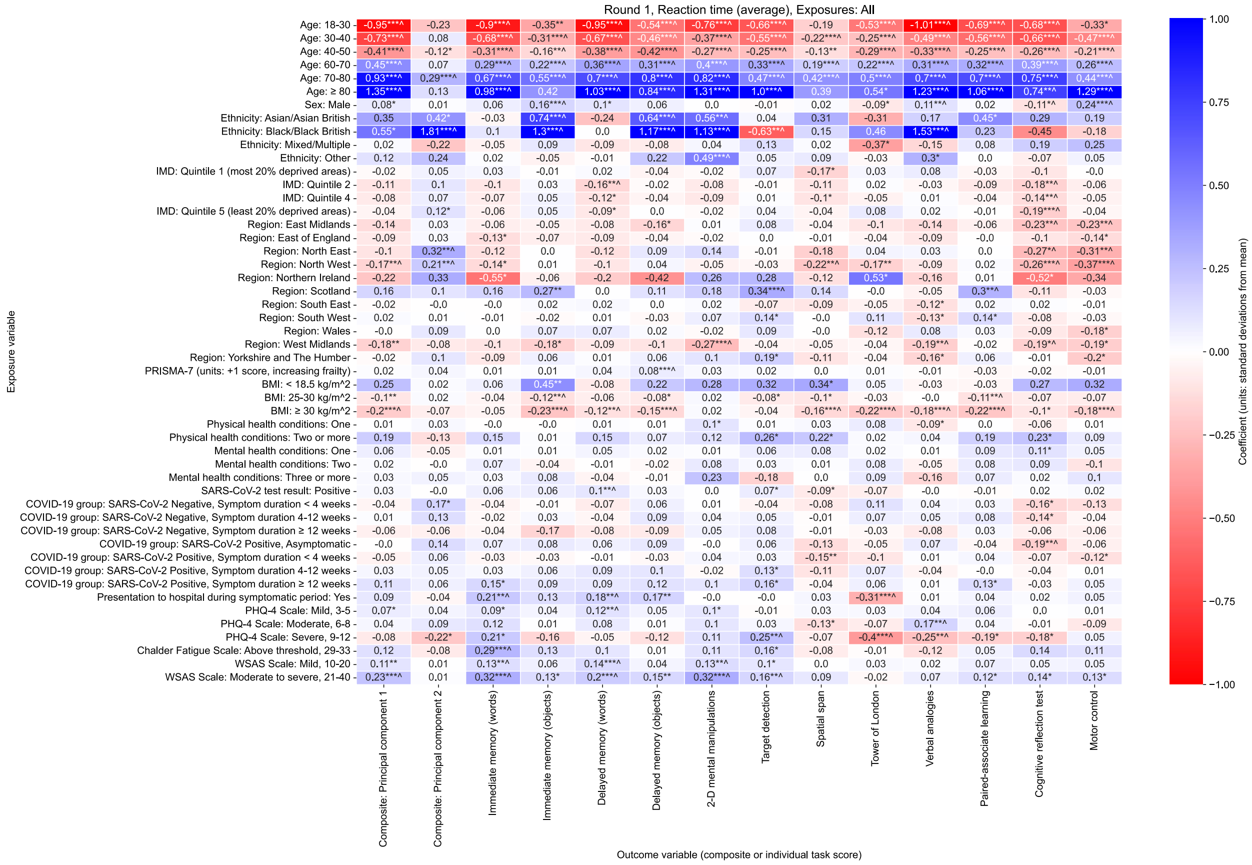


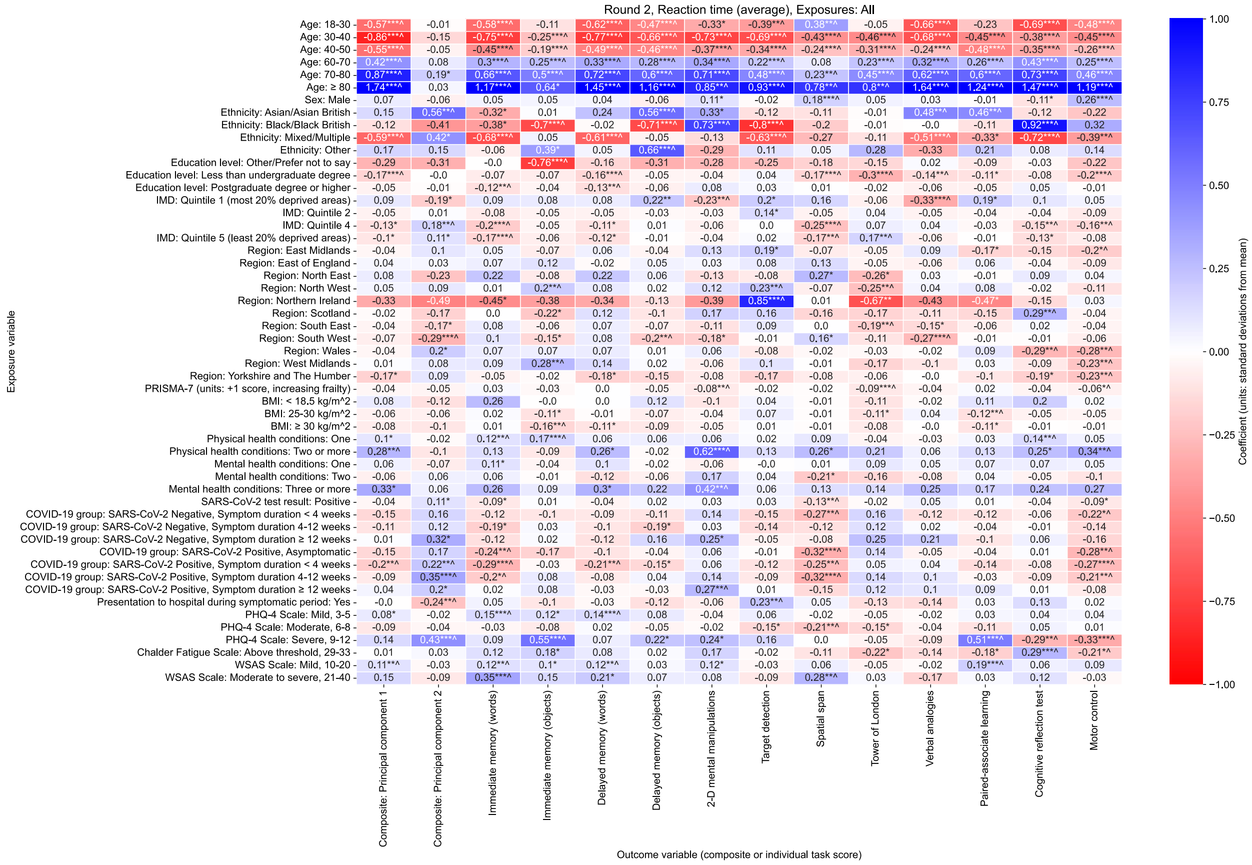


Figure S 10. **Associations with cognitive assessment average reaction time**. Standardised coefficients (number of standard deviations from mean) from multivariable ordinary least squares linear regression models testing association between various exposure variables and average reaction time during cognitive tasks as outcome variables. Results are from Round 1 and Round 2 of cognitive testing, among all individuals who completed either Round 1 or Round 2 of testing. Asterisk symbols represent uncorrected p-values, * < 0.05, ** < 0.01, *** < 0.001, while superscript caret symbol ^ illustrates $p < 0.05$ after adjustment for multiple testing. Results for each combination of both exposure variable and outcome variable presented originate from separate models that use distinct adjustment variable sets determined from the proposed DAG for cognitive performance.

#### S16. Cognitive accuracy – mediation models

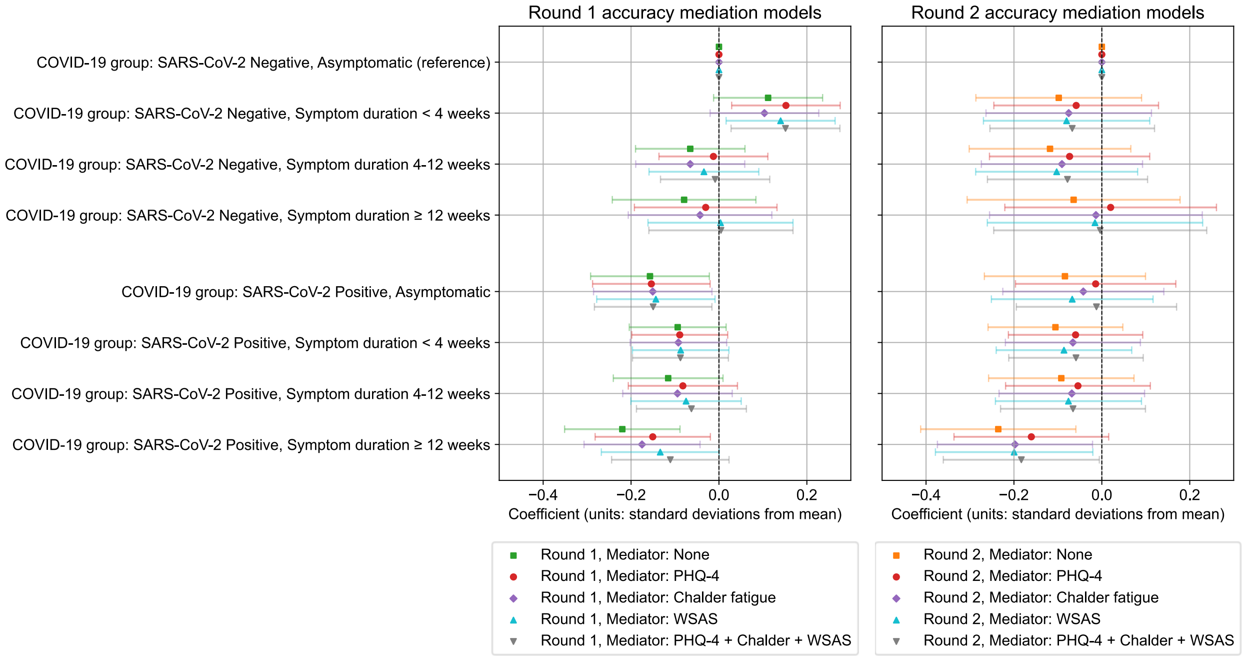


Figure S 11. **Mediation of the effect of COVID-19 group on cognitive accuracy by current symptoms.** Standardised coefficients (number of standard deviations from mean) with 95% confidence intervals from multivariable ordinary least squares linear regression models testing association between COVID-19 group and cognitive accuracy PCA 1st component standardised scores. Results are from Round 1 (left) and Round 2 (right) of cognitive testing, among all individuals who completed either round of testing. The following set of adjustment variables were used within each model: Age, BMI, Deprivation, Education (Round 2 models only), Ethnicity, Frailty (PRISMA-7), Mental health condition count, Physical health condition count, Presentation to hospital, Region, Sex.

#### S17. Correlation between self-reported COVID-19 recovery and symptom duration

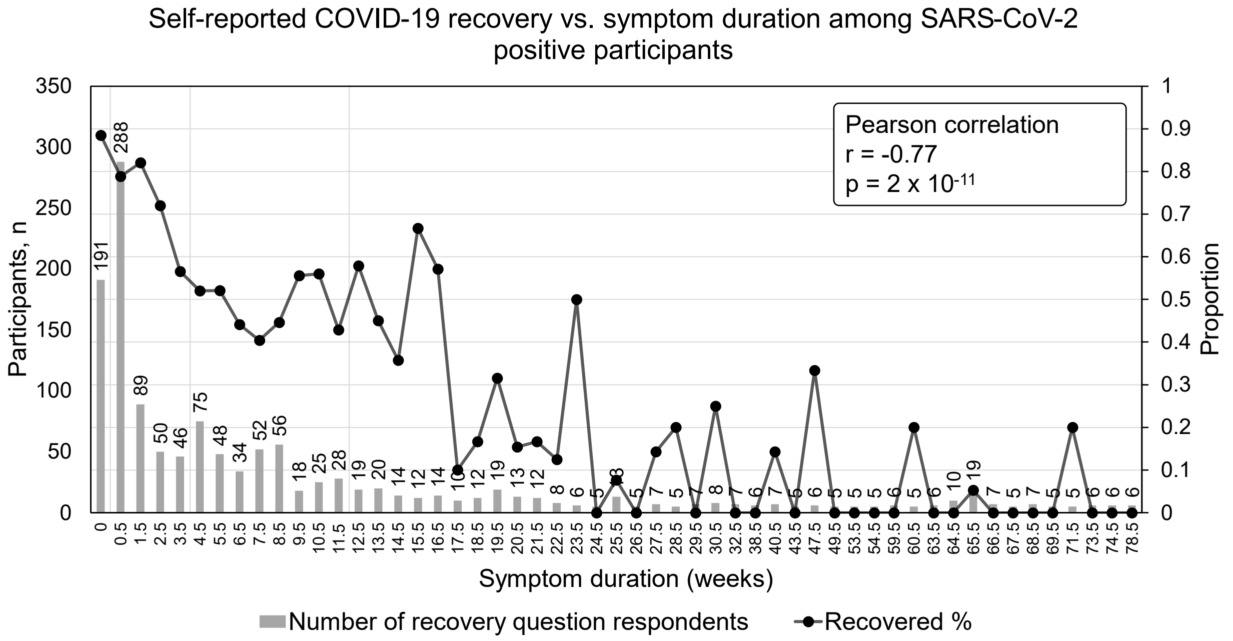


Figure S 12. **Correlation between self-reported COVID-19 recovery and COVID-19 symptom duration among SARS-CoV-2 positive participants**. (Left axis, grey bars) number of participants, SARS-CoV-2 positive at time of Round 1 of cognitive testing, who participated in Round 1 of cognitive testing and answered self-reported COVID-19 recovery survey question. (Right axis, black marker and line) proportion of participants who reported as “Yes, I am back to normal” to survey question “Thinking about the last or only episode of COVID-19 you have had, have you now recovered and are back to normal?”. SARS-CoV-2 positive participants of symptom duration is estimated at time of Round 1 of cognitive testing. Asymptomatic participants are shown with duration = 0, while symptomatic duration is aggregated into number of weeks, and shown at the mid-point of the given number of weeks, e.g., duration 1-6 days plotted at 0.5 weeks. Symptom duration in weeks with fewer than 5 participants were excluded from plot and Pearson correlation statistic calculation.
