## Supplementary material for "The effects of COVID-19 on cognitive performance in a community-based cohort: A COVID Symptom Study Biobank observational study": ICMJE disclosure forms: coi_disclosure_ErikaMolteni.pdf

### ICMJE DISCLOSURE FORM

| | |
|---|---|
| **Date:** | 11/23/2022 |
| **Your Name:** | Erika Molteni |
| **Manuscript Title:** | The effects of COVID-19 symptom duration, recovery, and infection timing on cognitive performance in a community-based cohort: A Covid Symptom Study Biobank observational study |
| **Manuscript Number (if known):** | Click or tap here to enter text. |

**Please place an "X" next to the following statement to indicate your agreement:**

☒ I certify that I have answered every question and have not altered the wording of any of the questions on this form.

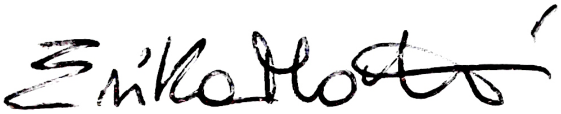
